# Modeling and Interpreting Patient Subgroups in Hospital Readmission: Visual Analytical Approach

**DOI:** 10.1101/2022.02.27.22271534

**Authors:** Suresh K. Bhavnani, Weibin Zhang, Shyam Visweswaran, Mukaila Raji, Yong-Fang Kuo

## Abstract

**Background:** A primary goal of precision medicine is to identify patient subgroups and infer their underlying disease processes, with the aim of designing targeted interventions. However, while several studies have identified patient subgroups, there is a considerable gap between the identification of patient subgroups, and their modeling and interpretation for clinical applications.

**Objectives:** To develop and evaluate a novel analytical framework for modeling and interpreting patient subgroups (MIPS) using a three-step modeling approach. (1) *Visual analytical* modeling to automatically identify patient subgroups and their co-occurring comorbidities, and determine their statistical significance and clinical interpretability. (2) *Classification* modeling to classify patients into subgroups and measure its accuracy. (3) *Prediction* modeling to predict a patient’s risk for an adverse outcome, and compare its accuracy with and without patient subgroup information.

**Methods:** The MIPS framework was developed using (1) bipartite networks to identify patient subgroups based on frequently co-occurring high-risk comorbidities; (2) multinomial logistic regression to classify patients into subgroups; and (3) hierarchical logistic regression to predict the risk of an adverse outcome using subgroup membership, compared to standard logistic regression without subgroup membership. The MIPS framework was evaluated on three hospital readmission conditions: chronic obstructive pulmonary disease (COPD), congestive heart failure (CHF), and total hip/knee arthroplasty (THA/TKA). For each condition, we extracted cases defined as patients readmitted within 30 days of hospital discharge, and controls defined as patients not readmitted within 90 days of discharge, matched by age, gender, race, and Medicaid eligibility (n[COPD]=29,016, n[CHF]=51,550, n[THA/TKA]=16,498).

**Results:** In each condition, the visual analytical model identified patient subgroups that were statistically significant (Q=0.17, 0.17, 0.31; *P*<.001, <.001, <.05), were significantly replicated (RI=0.92, 0.94, 0.89; *P*<.001, <.001, <.01), and were clinically meaningful to clinicians. (2) In each condition, the classification model had high accuracy in classifying patients into subgroups (mean accuracy=99.60%, 99.34%, 99.86%). (3) In two conditions (COPD, THA/TKA), the hierarchical prediction model had a small but statistically significant improvement in discriminating between the readmitted and not readmitted patients as measured by net reclassification improvement (NRI=.059, .11), but not as measured by the C-statistic or integrated discrimination improvement (IDI).

**Conclusions:** While the visual analytical models identified statistically and clinically significant patient subgroups, the results pinpoint the need to analyze subgroups at different levels of granularity for improving the interpretability of intra- and inter-cluster associations. The high accuracy of the classification models reflects the strong separation of the patient subgroups despite the size and density of the datasets. Finally, the small improvement in predictive accuracy suggests that comorbidities alone were not strong predictors for hospital readmission, and the need for more sophisticated subgroup modeling methods. Such advances could improve the interpretability and predictive accuracy of patient subgroup models for reducing the risk of hospital readmission and beyond.

## INTRODUCTION

### Overview

A wide range of studies [1–9] on topics ranging from molecular to environmental determinants of health have shown that most humans tend to share a subset of characteristics (e.g., comorbidities, symptoms, genetic variants), forming distinct patient subgroups. A primary goal of precision medicine is to identify such patient subgroups and infer their underlying disease processes to design interventions targeted to those processes [2, 10]. For example, recent studies in complex diseases such as breast cancer [3, 4], asthma [5–7] and COVID-19 [11] have revealed patient subgroups, each with different underlying mechanisms precipitating the disease, and therefore each requiring different interventions.

However, there is a considerable gap between the identification of patient subgroups, and their modeling and interpretation for clinical applications. To bridge this gap, we developed and evaluated a novel analytical framework called Modeling and Interpreting Patient Subgroups (MIPS) using a three-step modeling approach: (1) identification of patient subgroups, their frequently co-occurring characteristics, and their risk for adverse outcomes, (2) classification of a new patient into one or more subgroups, and (3) prediction of an adverse outcome for a new patient informed by subgroup membership. We evaluated MIPS on three datasets related to hospital readmission, which helped to pinpoint the strengths and limitations of MIPS. The results provide implications of MIPS for improving the interpretability of patient subgroups in large and dense datasets, and for the design of clinical decision support systems to prevent adverse outcomes such as hospital readmissions.

### Identification of Patient Subgroups

Patients have been divided into subgroups by using (a) investigator-selected variables such as race for developing hierarchical regression models [12], or assigning patients to different arms of a clinical trial, (b) existing classification systems such as the Medicare Severity-Diagnosis Related Group (MS-DRG) [13] to assign patients into a disease category for purposes of billing, and (c) computational methods such as classification [14–16] and clustering [5, 17] to discover patient subgroups from data (also referred to as *subtypes* or *phenotypes* depending on the condition and variables analyzed).

Several studies have used computational methods to identify patient subgroups, each with critical trade-offs. Some studies have used *combinatorial* approaches [18] (identify all pairs, all triples etc.), which are intuitive, but which can lead to a combinatorial explosion (e.g., enumerating combinations of the 31 Elixhauser comorbidities would lead to 2^31^ or 2147483648 combinations), with most combinations that do not incorporate the full range of symptoms (e.g., the most frequent pair of symptoms ignores what other symptoms exist in the profile of patients with that pair). Other studies have used *unipartite* clustering methods [16, 17] (clustering patients or comorbidities, but not both together) such as k-means, and hierarchical clustering; and dimensionality-reduction methods such as principal component analysis (PCA) to help identify clusters of frequently co-occurring comorbidities [18–24]. However, such methods have well-known limitations including the requirement of inputting user-selected parameters (e.g., similarity measures, and the number of expected clusters), in addition to the lack of a quantitative measure to describe the quality of the clustering (critical for measuring the statistical significance of the clustering). Furthermore, because these methods are unipartite, there is no agreed-upon method to identify the patient subgroup defined by a cluster of variables, and vice-versa.

More recently, bipartite network analysis [25] has been used to address the above limitations by automatically identifying *biclusters,* consisting of patients and characteristics simultaneously. This method takes as input any dataset such as patients and their comorbidities, and outputs a quantitative and visual description of biclusters (containing both patients subgroups and their frequently co-occurring comorbidities). The quantitative output generates the number, size, and statistical significance of the biclusters [26–28], and the visual output displays the quantitative information of the biclusters through a network visualization [29–31]. Bipartite network analysis therefore enables (1) the automatic identification of biclusters and their significance, and (2) the visualization of the biclusters critical for their clinical interpretability. Furthermore, the attributes of patients in a subgroup can be used to measure the subgroup risk for an adverse outcome, to develop classifiers for classifying a new patient into one or more of the subgroups, and to develop a predictive model that uses that subgroup membership for measuring the risk of an adverse outcome for the classified patient.

However, while several studies [11, 28, 32–38] have demonstrated the usefulness of bipartite networks for the identification and clinical interpretation of patient subgroups, there has been no systematic attempt to integrate them with classification and prediction modeling, a critical step towards clinical application. We therefore leveraged the advantages of bipartite network to develop the MIPS framework, with the goal of bridging the gap between the identification and interpretation of patient subgroups, and their future clinical application.

### The Need for Modeling and Interpreting Patient Subgroups in Hospital Readmission

An estimated one in five elderly patients (over 2.3 million Americans) is readmitted to a hospital within 30-days after being discharged [39]. While many readmissions are unavoidable, an estimated 75% of readmissions are unplanned and mostly preventable [40], imposing a significant burden in terms of mortality, morbidity, and resource consumption. Across all conditions, unplanned readmissions cost almost $17 billion annually in the US [40], making them an ineffective use of costly resources, and therefore closely scrutinized as a marker for the poor quality of care by organizations such as the Centers for Medicare & Medicaid Services (CMS) [41].

To address this epidemic of hospital readmission, CMS sponsored the development of models to predict the patient-specific risk of readmission in specific index conditions such as chronic obstructive pulmonary disease (COPD) [42], congestive heart failure (CHF) [43], and hip/knee arthroplasty (THA/TKA) [44]. The independent variables include prior comorbidities (as recorded in Medicare claims data), and demographics (age, gender, and race). This was motivated by numerous studies that have shown that almost two-thirds of older adults have two or more comorbid conditions, which have a heightened risk for adverse health outcomes such as hospital readmission [45]. However, although prior studies have shown the existence of subgroups among patients with hospital readmission [28], none of the CMS models incorporated patient subgroups. The identification and inclusion of patient subgroups could improve the accuracy of predicting hospital readmission for a patient, in addition to enabling the design of interventions targeted to each patient subgroup for reducing the risk of readmission. We therefore used the MIPS framework to model and interpret patient subgroups in hospital readmission, and tested its generality across three index conditions. Furthermore, to enable a head-to-head comparison with the existing CMS predictive models, we used the same independent variables in addition to patient subgroup membership, when developing our prediction models.

## METHOD

### Overview of MIPS

Fig. 1 provides a conceptual description of the data inputs and outputs from the three-step modeling in MIPS. As shown, the visual analytical model identifies the patient subgroups, and visualizes them through a network. The classification model determines subgroup membership for cases and controls. These subgroup memberships are then used to measure the risk of readmission within each subgroup based on its proportion of cases, and juxtaposed with the respective subgroup visualization to enable clinicians interpret the readmitted patient subgroups. Finally, the predictive model uses the subgroup membership assignment of cases and controls to determine the readmission risk of a patient. Appendix-1 (Table-1) provides a summary of the inputs, methods, and outputs from each model.

**Fig. 1.**
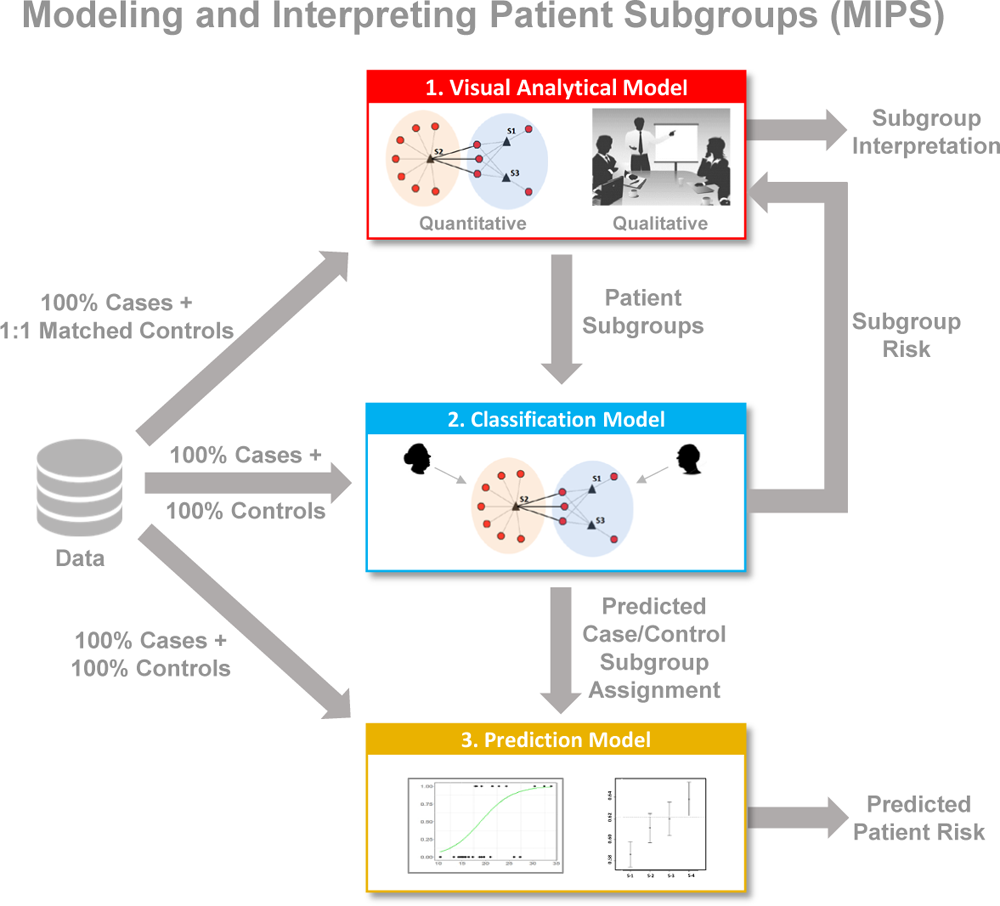
Inputs and outputs for the three-step modeling in MIPS consisting of the visual analytical model, the classification model, and the predictive model.

### Data Description

#### Study population

We analyzed patients hospitalized for chronic obstructive pulmonary disease (COPD), congestive heart failure (CHF), and total hip/knee arthroplasty (THA/TKA). We selected these three index conditions because: (a) hospitalizations for each of these conditions are highly prevalent in older adults [39]; (b) hospitals report very high variations in their readmission rates [39]; and (c) there exist well-tested readmission prediction models for each of these conditions that did not consider patient subgroups [42-44, 46, 47].

The data for these three index conditions were extracted from the Medicare insurance claims dataset. In 2019, Medicare provided health insurance to approximately 64.4 million Americans, of which 55.5 million were older Americans (>= to 65 years) [48]. Furthermore, 94% percent of non-institutionalized older Americans were covered by Medicare [49], with eligible claims received from 6,204 medical institutions across the US, and is therefore one of the few datasets that is highly representative of older Americans and their care.

For each index condition, we used the same inclusion and exclusion criteria that had been used to develop the CMS models, but with the most recent years (2013-2014) provided by Medicare when we started the project. We extracted all patients that were admitted to an acute care hospital on or after July 2013-August 2014 with a principal diagnosis of the index condition, were 66 years of age or older, and were enrolled in both Medicare parts A and B fee-for-service plans in the 6 months before admission. Furthermore, we excluded patients who were transferred from other facilities, died during the hospitalization, or transferred to another acute care hospital. Similar to the CMS models, we selected the first admission for patients with multiple admissions during the study period, and did not use Medicare Part D (related to prescription medications).

Appendix-2 describes (1) the International Classification of Diseases, Ninth Version codes (ICD-9) codes for each of the three index conditions selected for analysis, and (2) the inclusion and exclusion criteria used to extract cases and controls for COPD, CHF, and THA/TKA, the respective numbers of patients extracted at each step, and how we addressed the small incidence of missing data. Each modeling method used relevant subsets of the above data, as described in the section below on the analytical and evaluation approach.

#### Variables

The independent variables consisted of comorbidities, and patient demographics (age, gender, race). Comorbidities common in older adults were derived from three established comorbidity indices: Charlson Comorbidity Index (CCI) [50], Elixhauser Comorbidity Index (ECI) [51], and the Center for Medicare and Medicare Services Condition Categories (CMS-CC) used in the CMS readmission models [52] (the variables in the CMS models varied across the index conditions). As these indices had overlapping comorbidities, we derived a union of them, which was verified by the clinician stakeholders. They recommended that we also include the following additional variables as they were pertinent to each index condition: COPD (history of sleep apnea, mechanical ventilation); CHF (history of coronary artery bypass graft surgery); THA/TKA (congenital deformity of the hip joint, post-traumatic osteoarthritis). For each patient in our cohort, we extracted the above comorbidities and variables from the physicians, outpatient, and inpatient Medicare claims data in the 6 months before (to guard against miscoding), and on the day of the index admission. The dependent variable (outcome) was whether a patient with an index admission (COPD, CHF, THA/TKA) had an unplanned readmission to an acute-care hospital within 30 days of discharge, as was recorded in the MEDPAR file (inpatient claims) in the Medicare database.

### Analytical and Evaluation Approach

#### Visual Analytical Modeling

The goal of visual analytical modeling was to identify and interpret biclusters of readmitted patients (cases) consisting of patient subgroups and their most frequently co-occurring comorbidities. The data used to build the visual analytical model in each index condition consisted of randomly dividing 100% of the cases into a training (50%) and a replication (50%) dataset (we use the term *replication* to avoid confusion with the term *validation* typically used in classification and prediction models). For the feature selection, we extracted an equal number of 1:1 matched controls based on age, gender, and race/ethnicity, and Medicaid eligibility [53]. The above data were analyzed in each index condition using the following steps (Appendix-1 provides additional details for each step):

1. *Model Training*. To train the visual analytical model, we used feature selection for identifying the set of comorbidities that were univariably significant in both the training and replication datasets, and used bicluster modularity maximization [26, 27] for identifying the number, members, and significance of biclusters in the training dataset.
2. *Model Replication.* To test the replicability of the of biclusters, we repeated the above bicluster analysis on the replication dataset, and used the Rand Index (RI) [54] to measure the degree and significance of similarity in comorbidity co-occurrence between the two datasets.
3. *Model Interpretation.* To enable clinical interpretation of the patient subgroups, we used the *Fruchterman-Reingold* (FR) [29] and *ExplodeLayout* [30, 31] algorithms to visualize the network. Furthermore, based on a request from our clinician stakeholder team, for each bicluster we ranked and displayed the comorbidity labels with their univariable ORs for readmission (obtained from the feature selection above), and juxtaposed the readmission risk of the bicluster (obtained from the classification step discussed below) onto the network visualization. Clinician stakeholders were asked to use the visualization to interpret the patient subgroups, their mechanisms, and potential interventions to reduce the risk for readmission.

#### Classification Modeling

The goal of the classification modeling was to classify all cases and controls from the entire Medicare dataset into the biclusters identified from the visual analytical model. The resulting bicluster membership for all cases and controls were designed to (a) develop the predictive modeling described below, and (b) measure the risk of each subgroup to enable clinical interpretation of the patient sugroups. The training dataset in each condition consisted of a random sample of 75% cases with their subgroup membership (output of the visual analytical modeling), and an internal validation dataset consisting of randomly selected 25% of the cases (with subgroup membership used to validate the model). The above data were used to develop and use classification models in each index condition using the following steps (Appendix-1 provides additional details for each step):

1. *Model Training.* To train the classifier, we used multinomial logistic regression [16] with independent variables consisting of comorbidities (identified through the feature selection). Accuracy of the trained model was measured by calculating the percentage of times the model correctly classifed the cases into the subgroups, using the highest predicted probability across the subgroups.
2. *Model Internal Validation.* To internally validate the classifier, we randomly split the above data into the training (75%) and testing (25%) datasets, 1000 times. For each iteration, we trained a model using the training dataset, and measured its accuracy on the testing dataset. This was done by predicting the subgroup membership using the highest predicted probability among all the subgroups. The overall predicted accuracy was then estimated by calculating the mean accuracy across the 1000 models.
3. *Model Application.* To generate data for the visual analytical and prediction models, the above classifier was used to classify 100% cases and controls from our entire Medicare dataset (July 2013-August 2014). The resulting classified data were used to measure the risk of each subgroup risk (juxtaposed onto the network visualization to enable clinical interpretation), and to conduct the following prediction modeling.

#### Prediction Modeling

The goal of the prediction modeling was to predict the risk of readmission for a patient taking into consideration subgroup membership. The data used to build the prediction models consisted of 100% cases and 100% controls with subgroup membership generated from the above classification modeling. These data were randomly spilt into the training (75%) and validation (25%) datasets. The above data were used to train, internally validate, and compare prediction models in each index condition using the following steps (Appendix-1 provides additional details for each step):

1. *Model Training.* To train the prediction model, we used binary logistic regression for developing a Standard Model (without subgroup membership similar to the CMS models), and a Hierarchical Model (with subgroup membership). Independent variables for both models consisted of comorbidities (identified through the feature selection) and demographics, and the outcome was 30-day unplanned readmission (yes vs. no).
2. *Model Internal Validation.* To internally validate the models, we used the internal validation data set to measure discrimination (C-statistic), and calibration (calibration-in-the-large, and calibration slope).
3. *Model Comparisons.* To compare the accuracy of the Standard Model versus the Hierarchical Model, we used the chi-squared test to compare their C-statistics. Furthermore, to examine how the Standard Model performed on each subgroup, we measured the C-statistic of the Standard Model applied to each subgroup separately. Finally, because both of the above models used comorbidities selected through feature selection, they differed from the set of comorbidities used in the published CMS models. Therefore, to perform a head-to-head comparison with the published CMS models (COPD [42], CHF [43], and THA/TKA [44]), we developed a logistic regression model using the independent variables from the published CMS model (CMS Standard Model), and compared it to the same model but which also included subgroup membership (CMS Hierarchical Model). Similar to the above comparisons, we used the chi-squared test to compare the C-statistic of the CMS Standard Model versus the CMS Hierarchical Model, and additionally measured the differences between the models using Net Reclassification Improvement (NRI), Integrated Discrimination Improvement (IDI).

## RESULTS

### Data

Table 1 provides a summary of the number of cases and/or controls used to develop the three models in each condition.

**Table 1.** Training and replication/validation datasets used to develop the three models in each of the three index conditions.

| Model | Training | Replication/<br>Validation | Total |
| --- | --- | --- | --- |
| **Visual Analytical\*** |  |  |  |
| COPD (cases/controls) | 14,508/14,508 | 14,508/14,508 | 29,016/29,016 |
| CHF (cases/controls) | 25,775/25,775 | 25,775/25,775 | 51,550/51,550 |
| THA/TKA (cases/controls) | 8,249/8,249 | 8,249/8,249 | 16,498/16,498 |
| **Classification** |  |  |  |
| COPD (cases) | 10,842 | 3,615 | 14,457 |
| CHF (cases) | 19,254 | 6,418 | 25,672 |
| THA/TKA (cases) | 5,257 | 1,753 | 7,010 |
| **Prediction** |  |  |  |
| COPD (cases/controls) | 21,692/117,839 | 7,334/39,176 | 29,026/157,015 |
| CHF (cases/controls) | 38,728/183,093 | 12,845/61,095 | 51,573/244,188 |
| THA/TKA (cases/controls) | 12,376/255,203 | 41,44/85,049 | 16,520/340,252 |

\*The visual analytical models used 1:1 matched controls for the feature selection, and used only cases for the bipartite networks to analyze heterogeneity in readmission. The numbers shown for the visual analytical models are before removing patients with no comorbidities. The resulting cases-only datasets were used for the classification modelling as shown.

#### Visual Analytical Modeling

The visual analytical modeling of readmitted patients in all three index conditions produced statistically and clinically significant patient subgroups and their most frequently co-occurring comorbidities, which were significantly replicated. Results from each condition are described below.

##### COPD

The inclusion and exclusion selection criteria (see Appendix-2) resulted in a training dataset (n=14,508 matched case/control pairs, of which 51 patient pairs with no dropped comorbidities), and a replication dataset (n=14,508 matched case/control pairs, of which 51 patient pairs with no dropped comorbidities), matched by age, sex, race, and Medicaid eligibility (a proxy for economic status). The feature selection method (see Appendix-3) used 45 unique comorbidities identified from a union of the three comorbidity indices, plus 2 condition-specific comorbidities. Of these, 3 were removed because of <1% prevalence. Of the remaining, 30 survived the significance and replication testing with Bonferroni correction. The visual analytical model used these surviving comorbidities (d=30), and cases consisting of CHF readmitted patients with at least one of those comorbidities (n=14,457). As shown in Fig. 2, the bipartite network analysis identified 4 biclusters, each representing a subgroup of readmitted COPD patients and their most frequently co-occurring comorbidities. The biclustering had significant modularity (Q=0.17, z=7.3, *P*<.001), and significant replication (RI=0.92, z=11.62, *P*=<.001) of comorbidity co-occurrence. Furthermore, as requested by the clinician stakeholders, we juxtaposed a ranked list of comorbidities based on their ORs for readmission in each bicluster, in addition to the risk for each of the patient subgroups.

**Fig. 2.**
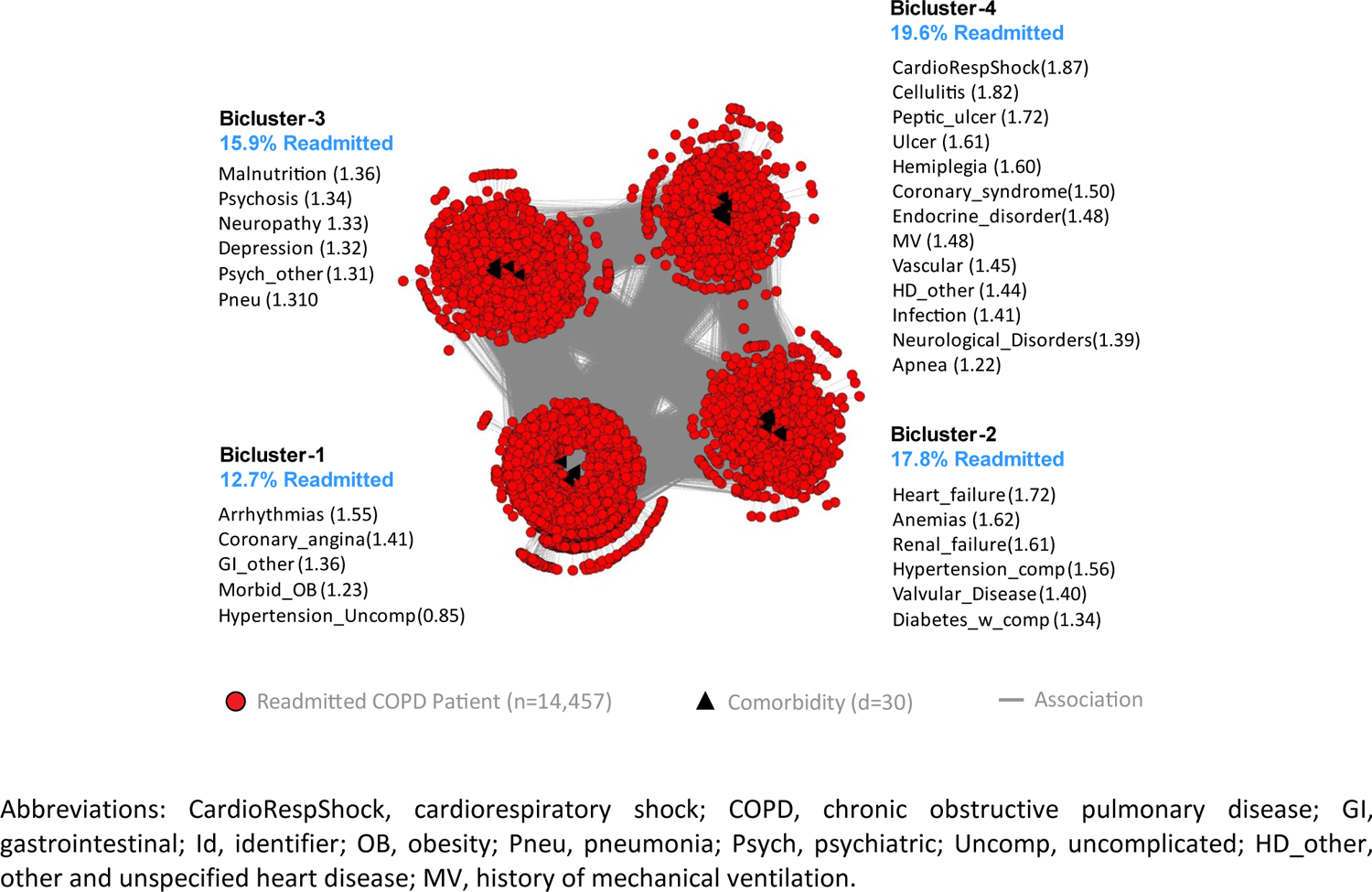
The COPD visual analytical model showing four biclusters consisting of patient subgroups and their most frequently co-occurring comorbidities (whose labels are ranked by their univariable ORs, shown within parentheses), and their risk of readmission (shown in blue text).

The pulmonologist inspected the visualization and noted that the readmission risk of the patient subgroups had a wide range (12.7% to 19.6%) with clinical (face) validity. Furthermore, the co-occurrence of comorbidities in each patient subgroup was clinically meaningful with interpretations for each subgroup. Subgroup-1 had a low disease burden with uncomplicated hypertension leading to the lowest risk (12.7%). This subgroup represented patients with early organ dysfunction and would benefit from using checklists such as regular monitoring of blood pressure in pre-discharge protocols to reduce the risk of readmission. Subgroup-3 had mainly psychosocial comorbidities, which could lead to aspiration precipitating pneumonia leading to an increased risk for readmission (15.9%). This subgroup would benefit from early consultation with specialists (e.g., psychiatrists, therapists, neurologists, and geriatricians) that had expertise in psycho-social comorbidities, with a focus on the early identification of aspiration risks and precautions. Subgroup-2 had diabetes with complications, renal failure and heart failure and therefore had higher disease burden leading to an increased risk for readmission (17.8%) compared to Subgroup-1. This subgroup had metabolic abnormalities with greater end-organ dysfunction and would therefore benefit from case management from advanced practice providers (e.g., nurse practitioners) with rigorous adherence to established guidelines to reduce the risk of readmission. Subgroup-4 had diseases with end-organ damage including gastro-intestinal disorders, and therefore had the highest disease burden and risk for readmission (19.6%). This subgroup would also benefit from case management with rigorous adherence to established guidelines to reduce the risk of readmission. Furthermore, as patients in this subgroup typically experience complications that could impair their ability to make medical decisions, they should be provided with early consultation with a palliative care team to ensure that care interventions align with patients’ preferences and values.

##### CHF

The inclusion and exclusion selection criteria (see Appendix-2) resulted in a training dataset (n=25,775 matched case/control pairs, of which 103 patient pairs with no dropped comorbidities) and a replication dataset (n=25,775 matched case/control pairs, of which 104 patient pairs with no dropped comorbidities), matched by age, sex, race, and Medicaid eligibility (a proxy for economic status). The feature selection method (see Appendix-3) used 42 unique comorbidities identified from a union of the three comorbidity indices, plus 1 condition-specific comorbidity. Of these, 1 comorbidity was removed because of <1% prevalence. Of those remaining, 37 survived the significance and replication testing with Bonferroni correction. The visual analytical model (Fig. 3) used these surviving comorbidities (d=37), and cases consisting of CHF readmitted patients with at least one of those comorbidities (n=25,672). As shown in Fig. 3, the bipartite network analysis of the CHF cases identified 4 biclusters, each representing a subgroup of readmitted CHF patients and their most frequently co-occurring comorbidities. The analysis revealed that the biclustering had significant modularity (Q=0.17, z=8.69, *P*<.001), and significant replication (RI=0.94, z=17.66, *P*<.001) of comorbidity co-occurrence. Furthermore, as requested by the clinicians, we juxtaposed a ranked list of comorbidities based on their ORs for readmission in each bicluster, in addition to the risk for each of the patient subgroups.

**Fig. 3.**
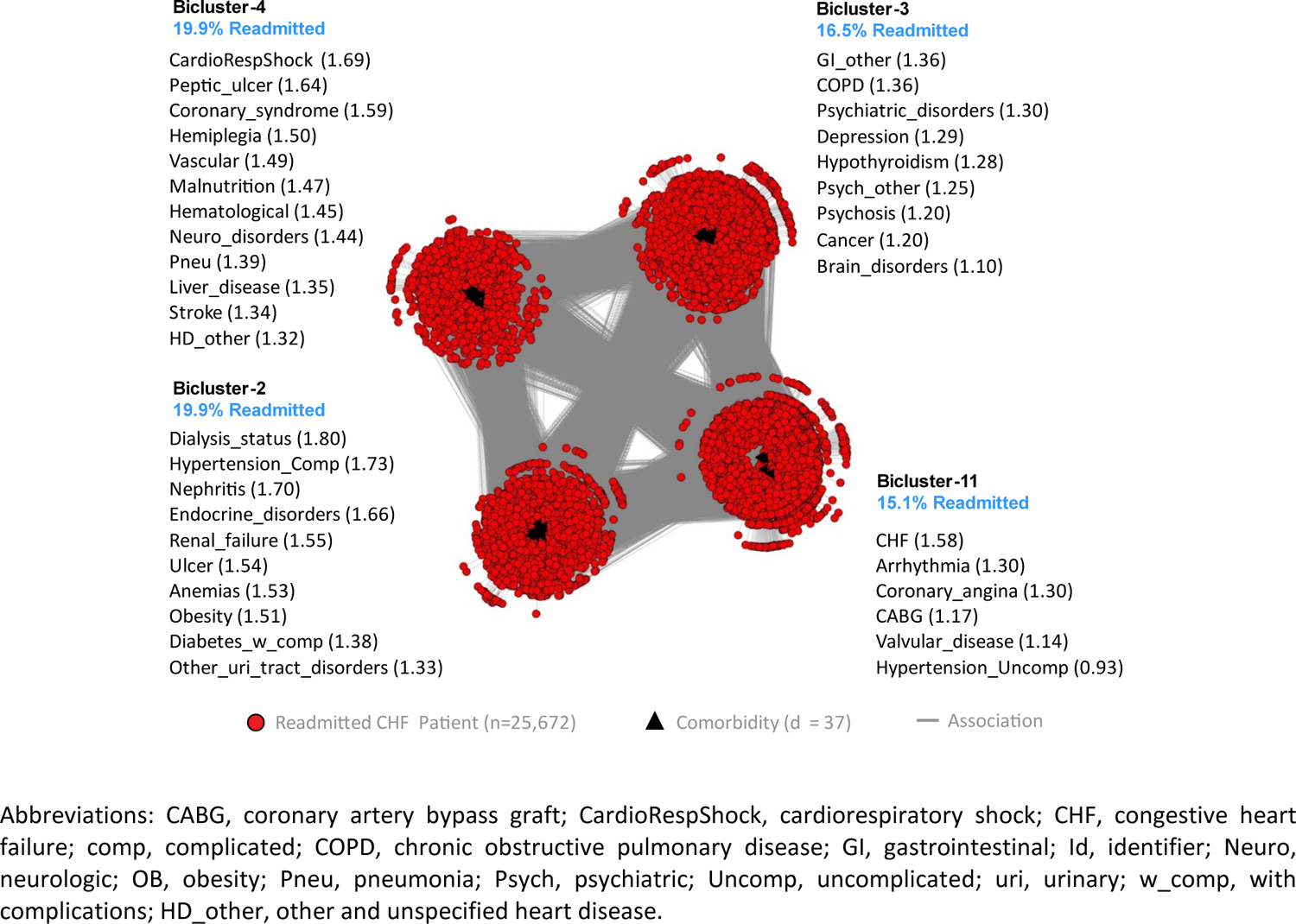
The CHF visual analytical model showing four biclusters consisting of patient subgroups and their most frequently co-occurring comorbidities (whose labels are ranked by their univariable ORs, shown within parentheses), and their risk of readmission (shown in blue text).

The geriatrician inspected the visualization and noted that the readmission risk of the patient subgroups, ranging from 15.1% to 19.9%, was wide with clinical (face) validity. Furthermore, the co-occurrence of comorbidities in each patient subgroup was clinically meaningful. Subgroup-1 had chronic but stable conditions, and therefore had the lowest risk for readmission (15.1%). Subgroup-3 had mainly psychosocial comorbidities, but were not as clinically unstable or fragile compared to subgroups 2 and 4, and therefore had medium risk (16.6%). Subgroup-2 had severe chronic conditions, making them clinically fragile (with potential benefits from early palliative and hospice care referrals), and were therefore at high risk for readmission if non-palliative approaches were used (19.9%). Subgroup-4 had severe acute conditions which were also clinically unstable, associated with substantial disability and care debility, and therefore at high risk for readmission and recurrent intensive care unit (ICU) use (19.9%).

##### THA/TKA

The inclusion and exclusion selection criteria (see Appendix-2) resulted in a training dataset (n=8,249 matched case/control pairs, of which 1239 patient pairs with no dropped comorbidities) and a replication dataset (n=8,249 matched case/control pairs, of which 1264 patient pairs with no dropped comorbidities), matched by age, sex, race, and Medicaid eligibility (a proxy for economic status). The feature selection (see Appendix-3) used 39 unique comorbidities identified from the three comorbidity indices plus 2 condition-specific comorbidities. Of these, 11 comorbidities were removed because of <1% prevalence. Of the remaining, 11 survived the significance and replication testing with Bonferroni correction. The visual analytical model (Fig. 4) used these surviving comorbidities (d=11), and cases consisting of readmitted patients with at least one of those comorbidities (n=7,010).

**Fig. 4.**
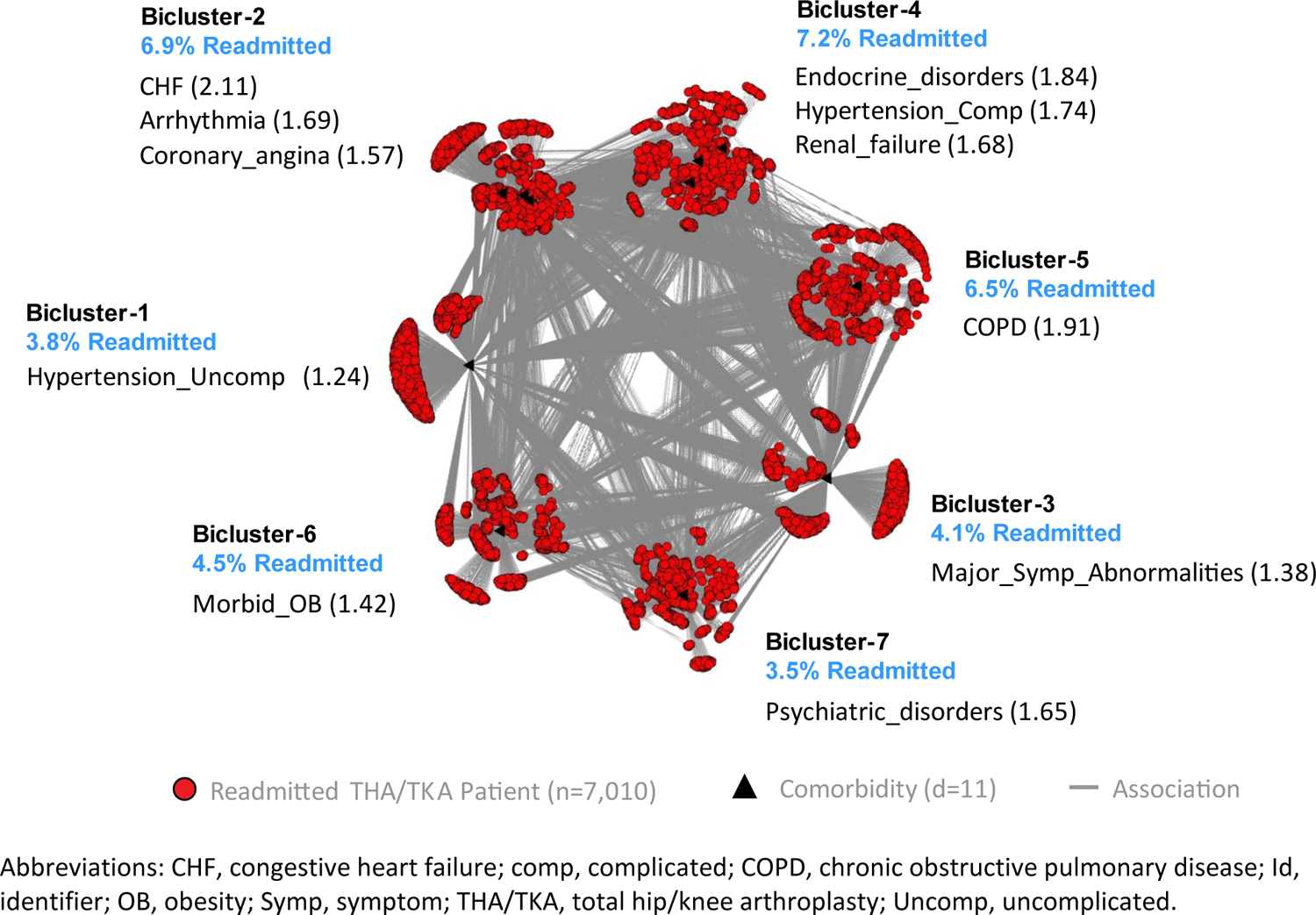
The THA/TKA visual analytical model showing four biclusters consisting of patient subgroups and their most frequently co-occurring comorbidities (whose labels are ranked by their univariable ORs, shown within parentheses), and their risk for readmission (shown in blue text).

As shown in Fig. 4, the bipartite network analysis of the THA/TKA cases identified 7 biclusters, each representing a subgroup of readmitted THA/TKA patients and their most frequently co-occurring comorbidities. The analysis revealed that the biclustering had significant modularity (Q=0.31, z=2.52, *P*=.011), and significant replication (RI=0.89, z=3.15, *P*=.002) of comorbidity co-occurrence. Furthermore, as requested by the clinician stakeholders, we juxtaposed a ranked list of comorbidities based on their ORs for readmission in each bicluster, in addition to the risk for each of the patient subgroups.

The geriatrician inspected the network and noted that TKA patients, in general, were healthier compared to THA patients, and therefore the network was difficult to interpret when the two index conditions were merged together. While our analysis was constrained because we were using the conditions as defined by CMS, these results nonetheless suggest that the interpretations did not suffer from a confirmation bias (manufactured interpretations to fit the results). However, he noted that the range of readmission risk had clinical (face) validity. Furthermore, subgroups 2, 4, and 5 had more severe comorbidities related to lung, heart, and kidney, and therefore had a higher risk for readmission compared to subgroups 1, 6, and 7 that had less severe comorbidities with a lower risk for readmission. In addition, subgroups 2, 5, 6 and 7 would benefit from chronic care case management from advanced practice providers (e.g., nurse practitioners). Finally, subgroups 2 and 5 could benefit from using well-established guidelines for CHF and COPD, subgroup 7 would benefit from mental health care and management of psycho-social comorbidities, and subgroup 6 would benefit from care for obesity and metabolic disease management.

### Classification Modeling

The classification model used multinomial logistic regression in each index condition (see Appendix-4 for the model coefficients) to predict the membership of patients using subgroups (identified from the above visual analytical models). The results revealed that in each index condition, the classification model had high accuracy in classifying all the cases in the full dataset (training dataset used in the visual analytical modeling). Similarly, the internal validation results using a 75%-25% split of the above dataset also had high classification accuracy (Table 2 with classification accuracy divided into quantiles). We report both results for each index condition:

**Table 2.** Internal validation results showing the percentage of COPD, CHF, and THA/TKA patients correctly-assigned to a subgroup by the classification models in each condition.

| Models | Quantiles |  |  |  |  | Summary |  |  |  |
| --- | --- | --- | --- | --- | --- | --- | --- | --- | --- |
|  | Q .025 | Q .25 | Q .50 | Q .75 | Q .975 | Min | Max | Mean | SD |
| **COPD** |  |  |  |  |  |  |  |  |  |
| Training (n=10842) | 99.90 | 100.00 | 100.00 | 100.00 | 100.00 | 99.70 | 100.00 | 100.00 | 0.02 |
| Testing (n=3615) | 99.30 | 99.40 | 99.60 | 99.60 | 99.80 | 99.10 | 100.00 | 99.60 | 0.15 |
| **CHF** |  |  |  |  |  |  |  |  |  |
| Training (n=19254) | 99.40 | 99.50 | 99.60 | 99.60 | 99.80 | 99.00 | 99.90 | 99.57 | 0.11 |
| Testing (n=6418) | 99.00 | 99.30 | 99.30 | 99.40 | 99.60 | 98.70 | 99.70 | 99.34 | 0.15 |
| **THA/TKA** |  |  |  |  |  |  |  |  |  |
| Training (n=5257) | 100.00 | 100.00 | 100.00 | 100.00 | 100.00 | 100.00 | 100.00 | 100.00 | 0.00 |
| Testing (n=1753) | 99.70 | 99.80 | 99.90 | 99.90 | 100.00 | 99.40 | 100.00 | 99.86 | 0.09 |

#### COPD

The model correctly predicted subgroup membership for 99.90% of the cases (14443/14457) in the full dataset. Furthermore, as shown in Table 2, the internal validation results revealed that the percentage of COPD cases correctly assigned to a subgroup in the testing dataset, ranged from 99.10% to 100.00%, with a median (Q.50) of 99.60%, and with 95% being in the range from 99.30% to 99.80%.

#### CHF

The model correctly predicted subgroup membership for 99.20% of the cases (25476/25672) in the full dataset. Furthermore, as shown in Table 3, the internal validation results revealed that the percentage of CHF cases correctly assigned to a subgroup in the testing dataset, ranged from 98.70% to 99.70%, with a median (Q.50) of 99.30%, and with 95% being in the range from 99.00% to 99.60%.

**Table 3.** Comparison of the CMS Standard Model with the CMS Hierarchical Model across the three index conditions based on NRI and IDI (* = significant at the .05 level).

| Model | NRI |  | IDI |
| --- | --- | --- | --- |
|  | Categorical (95% Interval) | Continuous (95% Interval) | IDI (95% Interval) |
| COPD | .023 (.012, .034)* | .059 (.034, .083)* | .0002 (-.0004, .0008) |
| CHF | -.010 (-.016, -.0004)* | -.038 (-.057, -.019)* | -.0006(-.0009, -.0003)* |
| THA/TKA | .022 (.012, .032)* | .111 (.080, .142)* | -.003(-.002, -.003)* |

#### THA/TKA

The model correctly predicted subgroup membership 100.00% of the cases (7010/7010) in the full dataset. Furthermore, as shown in Table 2, the internal validation results revealed that the percentage of CHF cases correctly assigned to a subgroup in the testing dataset, ranged from 99.40% to 100.00%, with a median (Q.50) of 99.90%, and with 95% being in the range from 99.70% to 100.00%.

### Application of the Classification Model to Generate Information for Other Models

The above classification model was used to classify 100% cases and 100% controls for use in the prediction model (described below). Furthermore, the proportion of cases and controls classified into each subgroup was used to calculate the risk of readmission for each subgroup (see Appendix-3). As this subgroup risk information was requested by the clinicians to improve interpretability of the visual analytical model, the values were juxtaposed next to the respective subgroups in the bipartite network visualizations (see blue text in Fig. 2-4).

### Prediction Modeling

For each of the three index conditions, we developed two binary logistic regression models to predict readmission, with comorbidities in addition to sex, age, and race: (1) **Standard Model** representing all patients without subgroup membership, similar to the CMS models; and (2) **Hierarchical Model** with an additional variable that adjusted for subgroup membership.

#### COPD

The inclusion and exclusion selection criteria (see Appendix-2) resulted in a cohort of 186,041 patients (29,026 cases and 157,015 controls). As shown in Fig. 5A, the Standard Model had a C-statistic of 0.624 (95% CI: 0.617-0.631) which was not significantly (*P*=.8578) different from the Hierarchical Model that had a C-statistic of 0.625 (95% CI: 0.618-0.632). The calibration plots revealed that both models had a slope close to 1, and an intercept close to 0 (see Appendix-5).

**Fig. 5.**
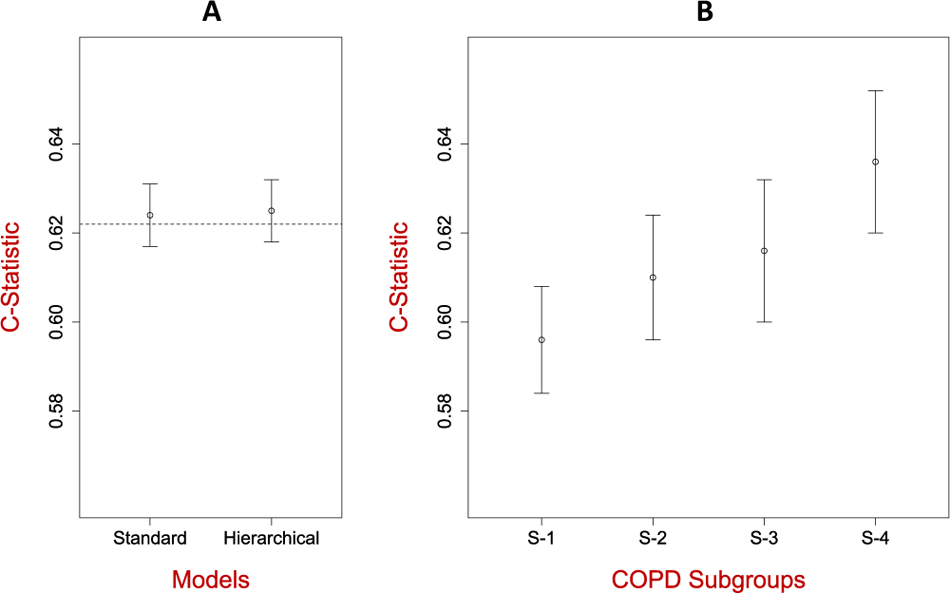
(A) Predictive accuracy of the Standard Model compared to the Hierarchical model in COPD, as measured by the C-Statistic. The C-statistic for the CMS Standard Model is shown as a dotted line. **(B)** Predictive accuracy of the Standard Model when applied separately to patients classified to each subgroup. S-1 has lower accuracy compared to S-3 and S-4. (C-statistics in A and B cannot be compared as they are based on models from different populations).

As shown in Fig. 5B, the Standard Model was used to measure the predictive accuracy of patients in each subgroup separately. The results showed that Subgroup-1 had a lower C-statistic compared to Subgroup-3 and Subgroup-4. While the C-statistics in Fig. 5A and Fig. 5B cannot be compared as they are based on models developed from different populations, these results reveal that the current CMS readmission model for CHF might be underperforming for one COPD patient subgroup, pinpointing which one might benefit by a Subgroup-Specific Model.

#### CHF

The inclusion and exclusion selection criteria (see Appendix-2) resulted in a cohort of 295,761 patients (51,573 cases and 244,188 controls). As shown in Fig. 6A, the Standard Model had a C-statistic of 0.600 (95% CI: 0.595-0.605), which was not significantly different (*P*=.2864) from the Hierarchical Model that also had a C-statistic of 0.600 (95% CI: 0.595-0.606). The calibration plots revealed that all models had a slope close to 1, and an intercept close to 0 (see Appendix-5).

**Fig. 6.**
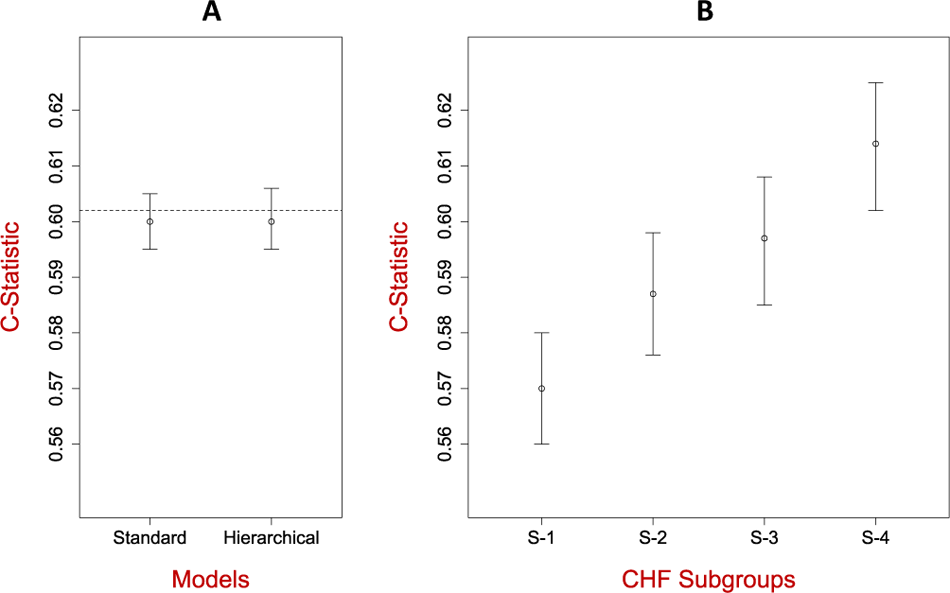
(A) Predictive accuracy of the Standard Model compared to the Hierarchical model in CHF as measured by the C-Statistic. The C-statistic for the CMS Standard Model is shown as a dotted line. **(B)** Predictive accuracy of the Standard Model when applied separately to patients classified to each subgroup. S-1 has lower accuracy compared to S-3 and S-4. (C-statistics in A and B cannot be compared as they are based on models from different populations).

As shown in Fig. 6B, the Standard Model was used to measure the predictive accuracy of patients in each subgroup separately. The results showed that Subgroup-1 had a lower C-statistic compared to Subgroup-4. While the C-statistics in Fig. 6A and Fig. 6B cannot be compared as they are based on models developed from different populations, but similar to the results in COPD, these results reveal that the current CMS readmission model for CHF might be underperforming for one CHF patient subgroup, pinpointing which one might benefit by a Subgroup-Specific Model.

#### THA/TKA

The application of the inclusion and exclusion selection criteria (see Appendix-2) resulted in a cohort of 356,772 patients (16,520 cases and 340,252 controls). As shown in Fig. 7A, the Standard Model had a C-statistic of 0.638 (95% CI: 0.629-0.646), which was not significantly different (*P*=.6817) from the Hierarchical Model that had a C-statistic of 0.638 (95% CI: 0.629-0.647). The calibration plots (see Appendix-5) revealed that both models had a slope close to 1, and an intercept close to 0 (see Appendix-5).

**Fig. 7.**
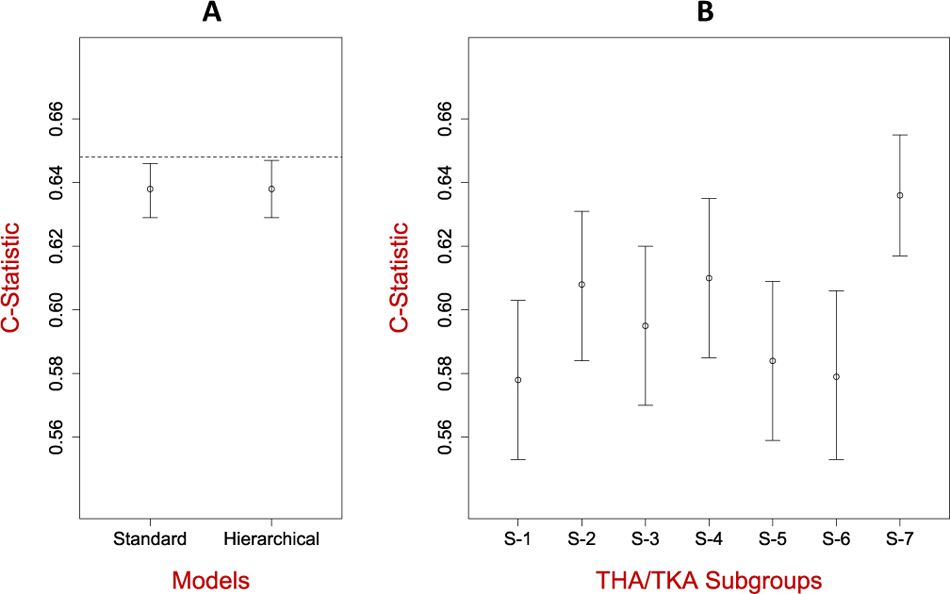
(A) Predictive accuracy of the Standard Model compared to the Hierarchical model in THA/TKA as measured by the C-Statistic. The C-statistic for the CMS Standard Model is shown as a dotted line. **(B)** Predictive accuracy of the Standard Model when applied separately to patients classified to each subgroup. S-1 has lower accuracy compared to S-7. (C-statistics in A and B cannot be compared as they are based on models developed from different populations).

As shown in Fig. 7B, the Standard Model was used to measure the predictive accuracy of patients in each subgroup separately. The results showed that Subgroup-1 had a lower C-statistic compared to Subgroup-4. Again, while the C-statistics in Fig. 7A and Fig. 7B cannot be compared as they are based on models developed from different populations, similar to the results in COPD, these results reveal that the current CMS readmission model for THA/TKA might be underperforming for 4 patient subgroups, pinpointing which ones might benefit by Subgroup-Specific Models.

### CMS Standard Model vs. CMS Hierarchical Model

Unlike the CMS published models, the above models used only the comorbidities that survived feature selection. Therefore, to perform a head-to-head comparison with the published CMS models, we also developed a CMS Standard Model (using the same variables from the published CMS model), and compared it to the corresponding CMS Hierarchical Model (with an additional variable for subgroup membership) in each condition. Similar to the models in Fig. 5-7, there were no significant differences in the C-statistics between the two modeling approaches in any condition (see Appendix-5). However, as shown in Table 3, the CMS Hierarchical Model for COPD had significantly higher NRI, but not significantly higher NDI compared to the CMS Standard Model; the CMS Hierarchical Model for CHF had a significantly lower NRI and IDI compared to the CMS Standard Model, and the CMS Hierarchical Model for THA/TKA had a significantly higher NDI and IDI compared to the CMS Standard Model. Furthermore, similar to the results in 6B-8B, when the CMS Standard Model was used to predict readmission separately in subgroups within each index condition, it identified subgroups that underperformed, pinpointing which ones might benefit by a Subgroup-Specific Model (See Appendix-5). In summary, the comparisons between the CMS Standard Models and the respective CMS Hierarchical Models showed that in two conditions (COPD and THA/TKA), there was a small but statistically significant improvement in discriminating between the readmitted and not readmitted patients as measured by NRI, but not as measured by the C-statistic or IDI, and that a subgroup in each index condition might be underperforming when using the CMS Standard Model.

## DISCUSSION

### Overview

Our overall approach of using the MIPS framework to identify patient subgroups through visual analytics, and using those subgroups to build classification and prediction models, revealed strengths and limitations for each modeling approach, and for our data source. This examination led to insights for developing future clinical decision support systems, and a methodological framework for improving the clinical interpretability of subgroup modeling results.

### Strengths and Limitations of Modeling Methods and Data Source

#### Visual Analytical Modeling

The results revealed three strengths of the visual analytical modeling: (1) the use of bipartite networks to simultaneously model patients and comorbidities, enabled the automatic identification of patient-comorbidity biclusters, and the integrated analysis of co-occurrence and risk; (2) the use of a bipartite modularity maximization algorithm to identify the biclusters enabled the measurement of the strength of the biclustering, critical for gauging its significance; and (3) the use of a graph representation enabled the results to be visualized through a network. Furthermore, the request from the clinician stakeholders to juxtapose the risk of each subgroup with their visualizations appeared to be driven by a need to reduce working memory loads (from having to remember that information spread over different outputs), which could have enhanced their ability to match bicluster patterns with chunks (previously-learned patterns of information) stored in long-term memory. The resulting visualizations enabled them to recognize subtypes based on co-occurring comorbidities in each subgroup, reason about the processes that precipitate readmission based on the risk of each subtype relative to the other subtypes, and propose interventions that were targeted to those subtypes and their risks. Finally, the fact that the geriatrician could not fully interpret the THA/TKA network because it mixed two fairly different conditions, suggests that the clinical interpretations were not the result of a *confirmation bias* (interpretations leaning towards fitting the results).

However, the results also revealed two limitations: (1) while modularity is estimated using a closed-form equation (formula), no closed-form equation exists to estimate the modularity variance, which is necessary to measure its significance. To estimate modularity variance, we therefore used a permutation test by generating 1000 random permutations of the data, and then compared the modularity generated from the real data to the mean modularity generated from the permuted data. Given the size of our datasets (ranging from 7K-25K patients), this computationally-expensive test took approximately 7 days to complete, despite the use of a dedicated server with multiple cores; and (2) while bicluster modularity was successful in identifying significant and meaningful patient-comorbidity biclusters, the visualizations themselves were extremely dense, and therefore potentially concealed patterns within and between the subgroups. Future research should explore a closed-form equation to estimate modularity variance, with the goal of accelerating the estimation of modularity significance, and more powerful analytical and visualization methods to reveal intra- and inter-cluster associations in large and dense networks.

#### Classification Modeling

The results revealed two strengths of the classification modeling: (1) the use of a simple multinomial classifier was adequate to predict with high accuracy to which subgroup a patient belonged; (2) because the model produced membership probabilities for each patient for each subgroup, the model captured the dense inter-cluster edges observed in the network visualization; and (3) the coefficients of the trained classifier could be inspected by an analyst making it more transparent (relative to most deep-learning classifiers which tend to be a black box).

However, because we dichotomized the classification probabilities into a single subgroup membership, our approach did not fully leverage the membership probabilities for modeling and visual interpretation. For example, some patients have high classification probabilities (representing strong membership) to a single subgroup (as shown by patients in the outer periphery of the biclusters with edges only within their bicluster), whereas others have equal probabilities to all subgroups (as shown in the inner periphery of the biclusters with edges going to multiple clusters). Future research should explore incorporating the probability of subgroup membership into the design of hierarchical models to improve predictive accuracy, and visualization methods to help clinicians interpret patients with different profiles of membership strength, with the goal of designing patient-specific interventions.

#### Predictive Modeling

The results revealed two strengths of the predictive modeling: (1) the use of the Standard Model to measure predictive accuracy across the subgroups helped to pinpoint which subgroups tend to have lower predictive accuracy compared to the rest, and therefore which of them could benefit from a more complex but accurate subgroup-specific model; and (2) despite the use of a simple Hierarchical Model with a dichotomized membership label for each patient, the predictive CMS models detected significant differences in the prediction accuracy as measured by NRI in two of the conditions, when compared to the CMS Standard Models. However, the results also revealed that the differences in predictive accuracy as measured by the C-statistic and NDI were small, suggesting that comorbidities on their own were potentially insufficient for accurately predicting readmission. Future research should explore the use of electronic health records, and the use of multiple subgroup-specific models targeted to each subgroup (enabling each model to have different slopes and intercepts), to potentially improve the predictive accuracy of the prediction models.

#### Data Source

The Medicare claims data had four key strengths: (1) scale of the datasets which enabled subgroup identification with sufficient statistical power; (2) spread of the data collected from across the US which enabled generalizability of the results; (3) data about older adults which enabled examination of subgroups in an underrepresented segment of the US population; and (4) data used by CMS to build predictive readmission models, which enabled a head-to-head comparison with the hierarchical modeling approach.

However, the data had two critical limitations. (1) As we compared our models with the CMS models, we had to use the same definition for controls (90 days with no readmission) that had been used, which introduced a selection bias that exaggerates the separation between cases and controls. Similarly, by excluding patients who died, this exclusion criterion potentially biased the results towards healthier patients. (2) Administrative data have known limitations such as the lack of comorbidity severity and test results, which could strongly impact the accuracy of predictive models. Future research should consider the use of national-level electronic health record (EHR) data such as those being assembled by the National COVID Cohort Collaborative (N3C) [55], and the TriNetX [56] initiatives, which could overcome the above limitations by providing laboratory values and comorbidity severity, but could also introduce new as yet unknown limitations.

#### Implications for Clinical Decision-Support that Leverage Patient Subgroups

While the focus of this project was to develop and evaluate the MIPS framework, its application to three index conditions coupled with extensive discussions with clinicians led to insights for designing a future clinical decision support system. Such a system could integrate outputs from all three models in MIPS. As we have shown, the visual analytical model automatically identified and visualized the patient subgroups, which enabled the clinicians to comprehend the co-occurrence and risk information in the visualization, reason about the processes that lead to readmission in each subgroup, and design targeted interventions. The classification model leveraged the observation that many patients have comorbidities in other biclusters (shown by a large number of edges between biclusters), and accordingly generated a membership probability of a patient belonging to each bicluster, from which the highest was chosen for bicluster membership. Finally, the predictive model predicts the risk for readmission for a patient, by using in the future the most accurate model designed for the bicluster to which the patient belongs.

The outputs from the above models could be integrated into a clinical decision support system to provide recommendations for a specific patient using the following algorithm: (1) use the classifier to generate the membership probability (MP) of a new patient belonging to each subgroup; (2) multiply the MP in each subgroup with the patient’s risk (R) for readmission provided by the predictive model for that subgroup, to generate an importance score [IS = *f*(MP) X *g*(R)] for the respective intervention; (3) rank the subgroups and their respective interventions using IS; and (4) use the ranking to display in descending order, the subgroup comorbidity profiles along with their respective potential mechanisms, recommended treatments, and the respective IS. Such model-based information, displayed through a user-friendly interface, could enable a clinician to rapidly scan the ranked list to (a) determine why a specific patient’s profile fits into one or more subgroups, (b) review the potential mechanisms and interventions ranked by their importance, and (c) use the combined information to design a treatment that is customized for the real-world context of the patient. Consequently, such a clinical decision support system could not only provide a quantitative ranking of membership to different subgroups, and the importance score for the associated interventions, but also enable the clinician to understand the rationale underlying those recommendations, making the system interpretable and explainable. Comparative evaluation of such a system to standard care could determine its clinical efficacy.

#### Implications for Analytical Granularity to Enhance the Interpretability of Patient Subgroups

While the visual analytical model enabled the clinicians to interpret the patient subgroups, they were unable to interpret the associations within and between the subgroups due to the large number of nodes in each bicluster and the dense edges between them. Several network filtering methods [57, 58] have been developed to “thin out” such dense networks such as by dropping or bundling nodes and edges based on user-defined criteria, to improve visual interpretation. However, such filtering could bias the results, or modify the clusters resulting from the reduced data.

An alternate approach that preserves the full dataset leverages the notion of analytical granularity, where the data is progressively analyzed at different levels. For example, we have analyzed COVID-19 patients [11] at the cohort, subgroup, and patient levels, and we are currently using the same approach to examine symptom co-occurrence and risk at each level in Long COVID patients. Our preliminary results suggest that analyzing data at different levels of granularity enables clinicians to progressively interpret patterns such as within and between subgroups, in addition to guiding the systematic development of new algorithms. For example, at the subgroup level, we have designed an algorithm that identifies which patient subgroups have a significantly higher probability for having characteristics that are clustered in another subgroup, providing critical information to clinicians about how to design interventions for such overlapping subgroups; at the patient level, we have identified patients that are outliers to their subgroups based on their pattern of characteristics inside and outside their subgroup. Such patient outliers could be flagged to examine if they need individualized interventions versus those recommended for the rest of their subgroup. Such analytical granularity could therefore inform the design of interventions by clinicians, in addition to the design of decision support systems that provide targeted and interpretable recommendations to physicians, who can then customize them to fit the real-world context of a patient.

#### Implications of the MIPS Framework for Precision Medicine

While we have demonstrated the application of the MIPS framework across multiple readmission conditions, its architecture has three properties that should enable its generalizability across other medical conditions. First, as shown in Fig. 1, the framework is *modular* with explicit inputs and outputs, enabling the use of other methods at each of the three modeling steps. For example, the framework could use other biclustering (e.g., Non-negative Matrix Factorization [59]), classification (e.g., deep learning [60]), and prediction methods (e.g., subgroup-specific modeling [16]). Second, the framework is *extensible,* enabling an elaboration of the methods at each modeling step to improve the analysis and interpretation of subgroups. For example, as discussed above, the analytical granularity at the cohort, subgroup, and patient levels could improve the interpretability of subgroups in large and dense datasets. Third, the framework is *integrative* as it systematically combines the strengths of machine learning, statistical, and precision medicine approaches. For example, the visual analytical modeling leverages search algorithms to discover co-occurrence in large datasets; the classification and prediction modeling leverages probability theory to measure the risk of co-occurrence patterns; and clinicians leverage medical knowledge and human cognition to interpret patterns of co-occurrence and risk for designing precision-medicine interventions. Such integration of different models and their interpretation operationalizes *team-centered informatics* [61] designed to facilitate data scientists, biostatisticians, and clinicians in multidisciplinary translational teams [62] to work more effectively across disciplinary boundaries, with the goal of designing interventions for precision medicine. Our current research tests the generality of the MIPS framework in other conditions such as Long COVID and Post-Stroke Depression, with the goal of designing and evaluating precision medicine interventions targeted to patient subgroups.

## CONCLUSIONS

Although several studies have identified patient subgroups in different conditions, there is a considerable gap between the identification of subgroups, and their modeling and interpretation for clinical applications. Here we developed MIPS, a novel analytical framework to bridge that gap using a three-step modeling approach. A visual analytical method automatically identified statistically significant and replicated patient subgroups and their frequently co-occurring comorbidities. Next, a multinomial logistic regression classifier had high accuracy in correctly classifying patients into the patient subgroups identified by the visual analytical model. Finally, despite using a simple hierarchical logistic regression model to incorporate subgroup information, the predictive models had a statistically significant improvement in discriminating between the readmitted and not readmitted patients in two of the three readmission conditions, and additional analysis pinpointed for which patient subgroups the current CMS model might be underperforming. Furthermore, the integration of the three models helped to (1) elucidate the data input and output dependencies among the models enabling clinicians to interpret the patient subgroups, reason about mechanisms precipitating hospital readmission, and design targeted interventions, and (2) provide a generalizable framework for the development of future clinical decision support systems that integrate outputs from each of the three modeling approaches.

However, evaluation MIPS across three readmission index conditions also helped to identify limitations of the models and the data. The visual analytical model was too dense to enable the clinicians to interpret the associations within and between the subgroups, and the absence of a closed-form equation to measure modularity variance required a computationally-expensive process to measure the significance of the biclustering. Furthermore, the small improvement in predictive accuracy suggested that comorbidities on their own were insufficient for predicting hospital readmission.

By leveraging the modular and extensible nature of the MIPs framework, future research should address the above limitations by developing more powerful algorithms which analyze subgroups at different levels of granularity to improve the interpretability of intra- and inter-cluster associations, and the evaluation of subgroup-specific models to predict outcomes. Furthermore, EHR data made available through national-level data initiatives such as N3C and TriNetX now provide access to critical variables including laboratory results and comorbidity severity, which should lead to higher predictive power for predicting adverse outcomes. Finally, extensive discussions with clinicians have confirmed the need for decision support systems which integrate outputs from the three models to provide for a specific patient, predicted subgroup memberships, ranked interventions, along with associated subgroup profiles and mechanisms. Such interpretable and explainable systems could enable clinicians to use patient subgroup information for informing the design of precision medicine interventions, with the goal of reducing adverse outcomes such as unplanned hospital readmissions and beyond.

## Data Availability

All data used in this study were available from the Centers of Medicare and Medicaid Services (CMS) after application with a fee, and signing a data use agreement (DUA) to analyze the deidentified data.

## AKNOWLEDGEMENTS

We thank Tianlong Chen, Clark Andersen, Yu-Li Lin, and Emmanuel Santillana for helping to conduct the analyses. This study was supported in part by the Patient-Centered Outcomes Research Institute (ME-1511-33194), the Clinical and Translational Science Award (UL1 TR001439) from the National Center for Advancing Translational Sciences at the National Institutes of Health, the UTMB Claude D Pepper Older Americans Independence Center funded by the National Institute of Aging at the National Institutes of Health (P30 AG024832), MD Anderson Cancer Center, and by the National Library of Medicine (R01 LM012095) at the National Institutes of Health. The content is solely the responsibility of the authors, and does not necessarily represent the official views of the Patient-Centered Outcomes Research Institute, or the National Institutes of Health. The Medicare data were analyzed using a CMS data-use agreement (CMS DUA RSCH-2017-51404).

## APPENDIX-1 Analytical Methods for the MIPS Framework

**Table 1.** Inputs used to train and replicate/validate the three models, and the analytical outputs they produced.

| Model | Inputs | Outputs |
| --- | --- | --- |
| **1. Visual Analytical Model**<br>(Bipartite Network Analysis) | • **Training Dataset:** 50% random sample of 100% cases, and an equal number of 1:1 matched controls (used only for feature selection)<br>• **Replication Dataset:** 50% random sample of 100% cases and equal number of 1:1 matched controls | • **Model Training**<br><u>Feature Selection:</u> Set of comorbidities univariably significant in both the training and replication datasets<br><u>Biclustering:</u> Modularity (degree of biclusteredness) and its significance, number of biclusters (subgroups), and their patient and comorbidity members in the training and replication datasets<br>• **Model Replication**<br><u>Comorbidity Co-Occurrence:</u> Rand Index (degree of replication), and its significance to measure replicability of comorbidity co-occurrence<br>• **Model Interpretation**<br><u>Visualization:</u> Layout of the bipartite network juxtaposed with risk of individual comorbidities and subgroups<br><u>Clinical Significance:</u> Interpretation by clinicians for face validity of patient subgroups based on comorbidity co-occurrence, leading to inference of mechanisms precipitating readmission, and interventions |
| **2. Classification Model**<br>(Multinomial Logistic Regression) | • **Training Dataset:** Random sample of 75% cases, with bicluster membership<br>• **Internal Validation Dataset:** Random sample of 25% of cases (with subgroup membership used to validate the model) | • **Model Training**<br><u>Subgroup Membership:</u> Probability of membership of each case to each subgroup (soft labels), with the highest used to determine subgroup membership (hard labels)<br>• **Model Internal Validation**<br><u>Internal Validation:</u> Accuracy of classification model based on hard labels<br>• **Model Application**<br><u>Classification:</u> Subgroup classification of 100% cases and 100% controls<br><u>Subgroup Risk:</u> Proportion of cases in each subgroup |
| **3. Prediction Model** (Binary Logistic Regression, and Hierarchical Binary Logistic Regression) | • **Training Dataset:** Random samples of 75% of 100% cases and controls, with subgroup membership<br>• **Internal Validation Dataset:** Random sample of 25% of cases and controls (with case/control labels used to validate the model) | • **Model Training**<br><u>Predicted Risk:</u> Each patient's probability of being readmitted.<br>• **Model Internal Validation**<br><u>Internal Validation:</u> C-statistic (discrimination), and calibration-in-the-large and calibration slope (calibration)<br>• **Model Comparison**<br><u>Accuracy:</u> Net Reclassification Improvement (NRI) and Integrated Discrimination Improvement (IDI) |

### Visual Analytical Modeling

The data used to build the visual analytical model consisted of 100% cases, and an equal number of 1:1 matched controls extracted by randomly selecting a control without replacement to match each case based on age, gender, race/ethnicity, and Medicaid eligibility [1]. The resulting dataset was divided randomly into a training (50%) and replication (50%) dataset (we use the term *replication* to avoid confusion with the term *validation* typically used in classification and prediction models). We used a bipartite network to model the cases (30-day readmitted patients) and significant comorbidities in each index condition using the following steps:

A. *Model Training.* The training of the bicluster network model consisted of the following two steps:
  I. *Feature Selection.* Given the large number of patients and comorbidities in the dataset, we used feature selection to identify comorbidities with the strongest signal and therefore interpretability for readmission using the following steps: (1) excluded comorbidities with prevalence less than 1% (as is commonly done in studies to reduce noise [2]); (2) selected significant comorbidities in the training dataset based on a 2-way interaction test using odds ratio (OR) with directionality, and correcting for multiple testing using Bonferroni, and (3) tested the surviving comorbidities for replication in the replication dataset, and selected those that were significant in both datasets. Appendix-2 shows the number of comorbidities, and variables that were included in the analysis for each of the three index conditions. The above feature selection generated a single set of significant and replicated comorbidities used for the following bipartite network analysis.
  II. *Biclustering.* We used bipartite networks analysis [3] on the training dataset to analyze heterogeneity in readmission using the following steps: (1) Removed all cases that did not have any comorbidities (as the modularity maximization algorithm will trivially put disconnected nodes into a separate cluster). (2) Represented the cases (30-day readmitted patients in the training dataset) and their significant and replicated comorbidities (selected in Step A) as a bipartite network. As shown in Fig. 1, the nodes represented cases (circles) or comorbidities (triangles), and edges (lines) represented which case had which comorbidity. (3) Used a bipartite modularity maximization algorithm [4–6], to identify the number of biclusters, their members, and degree of biclusteredness of the network using modularity. Modularity is defined as the fraction of edges falling within a cluster, minus the expected fraction of such edges in a network of the same size with randomly assigned edges. Modularity ranges from −0.5 to +1, with values >0 indicating biclustering that is higher than can be expected by chance. We used the bipartite version of modularity to find biclusters in the network. (4) Measured the significance of the bicluster modularity by comparing it to a distribution of the same quantity generated from 1000 random permutations of the network, by preserving the network size (number of nodes) and the network density (number of edges).
B. *Model Replication.* Repeated the above biclustering steps 1-4 to identify subgroups in the replication dataset, and compared the comorbidity co-occurrence in the training dataset, to that in the replication dataset using the Rand index (RI) [7]. RI measures the proportion of comorbidity pairs that co-occurred and did not co-occur in a cluster in the training and replication datasets (where 0=no inter-network cluster similarity, and 1=total inter-network cluster similarity). The significance of RI was measured by comparing it to a distribution of the same quantity generated from 1000 random permutations of the training and replication networks. All tests of statistical significance in Steps A and B were 2-sided.
C. *Model Interpretation.* The model interpretation consisted of the following steps:
  I. *Visualization.* We used the following steps to visualize the network generated from the training dataset. (1) Used *Fruchterman-Reingold* (FR) [8], a force-directed algorithm to lay out the bipartite network. This layout algorithm pulls together nodes that are strongly connected, and pushes apart nodes that are not. This results in nodes with a similar pattern of connections to be placed close to each other in Euclidean space, and those that are dissimilar are pushed apart. (2) As the FR algorithm often cannot entirely separate clusters in large and dense networks, the network layout needs to be visually enhanced before it is interpretable by clinician stakeholders. Therefore, we used the *ExplodeLayout* algorithm [9, 10] to separate the biclusters to reduce their visual overlap. This algorithm preserves the distances of nodes within a bicluster, but increases the distance of nodes between clusters to improve interpretability. (3) Juxtaposed the risk of readmission with the network visualization (in response to a request from the clinical stakeholders). This was done by (a) displaying comorbidity labels with their univariable ORs for readmission (measured in Step A) ranked by their odds ratios (ORs) for each subgroup, and (b) measuring the readmission risk for each patient subgroup based on the full case-control population (explained in more detail in the section on classification modeling), and juxtaposing it with the respective subgroup.
  II. *Clinical Interpretation.* We used the following steps to solicit clinical interpretations of the above bipartite network. (1) Recruited a pulmonologist specializing in COPD and hospital readmission to interpret the COPD results, and a geriatrician with expertise in treating older adults in CHF and THA/TKA to interpret the respective results. (2) Requested each clinician stakeholder to interpret the patient subgroups, their mechanisms, and potential interventions to reduce the risk for readmission.

**Fig. 1.**
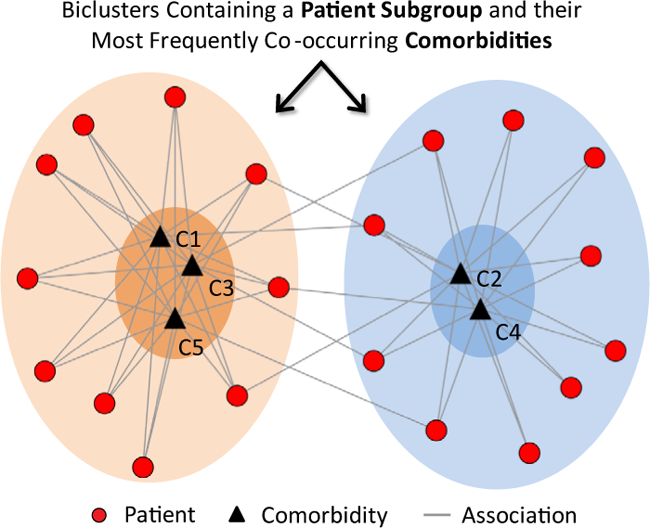
A bipartite network showing patient subgroups and their most frequently co-occurring comorbidities.

### Classification Modeling

As shown in the bipartite network example in Fig. 1, the biclusters identified through the modularity maximization algorithm contain patient subgroups and their *most frequently* co-occurring comorbidities with respect to other patients in the network. However, there are often many edges between biclusters, revealing that many patients within a bicluster have comorbidities that exist in other biclusters. As is true for most partitioning cluster methods, including modularity, membership of a new patient to each bicluster is therefore *probabilistic*. The classification of a patient into a cluster is therefore not defined by the *inclusion or exclusion of comorbidities* (e.g., hypertension and diabetes), but rather by the *probability* of being in a patient subgroup. Patients are therefore similar or different, not just in a handful of carefully-selected comorbidities while ignoring others, but based on *all* of their recorded comorbidities. This overall profile of patients reflects the reality of comorbid conditions.

To model the above complexity, we used multinomial logistic regression [11] to develop classification models in each index condition. This approach has the advantage of generating probabilities (“soft labels”) for a patient to belong to each patient subgroup. The models were trained, internally validated, and then applied to generate information for the other two modeling methods, as described below:

A. *Model Training.* The data used to build the classification model consisted of the training dataset and subgroup membership from the visual analytical model. We trained a multinomial logistic regression model using the above data, with independent variables that included comorbidities identified through feature selection done for the visual analytical modeling. Accuracy of the trained model was measured by calculating the percentage of times the model correctly classifed the cases into the subgroups, using the highest predicted probability across the subgroups (“hard labels”).
B. *Model Internal Validation.* To internally validate the classifier, we randomly split the above data into training (75%) and testing (25%) datasets, 1000 times. For each iteration, we trained a model using the training dataset, and measured its accuracy on the testing dataset. This was done by predicting the subgroup membership using the highest predicted probability among all the subgroups. The overall predicted accuracy was then estimated by calculating the mean accuracy across the 1000 models.
C. *Model Application.* Using the 100% cases, in addition to the 100% controls from July 2013-August 2014 (representing the entire Medicare population of each index condtion from those years), we generated the following two types of information for use in the other models. (1) Used the classifier trained in Step A above, to classify 100% cases and 100% controls into a subgroup. This information was used by the subsequent predictive modeling. (2) While the visual analytical model used the 1:1 matched controls for feature selection, this cohort did not represent the entire population. Therefore, to accurately measure the subgroup risk, we used the entire case-control population classified into the subgroups (as described in the above step), and measured the proportion of cases in each subgroup. Furthermore, as requested by the clinicians, we juxtaposed these subgroup risks next to the respective subgroups in the bipartite network visualization, to improve their interpretability.

### Predictive Modeling

The data used to build the predictive models consisted of 100% cases and 100% controls, in addition to their subgroup membership generated from the above classification models. These data were randomly spilt into a training (75%) and validation (25%) dataset. The predictive models were trained, internally validated, and compared for predictive accuracy, as described below:

A. *Model Training.* We used the training dataset to train a Standard Model (binary logistic regression without subgroup membership similar to the CMS models), and a Hierarchical Model (binary logistic regression with subgroup membership), with 30-day unplanned readmission (yes vs. no) as the outcome. Independent variables for both models included comorbidities identified through the feature selection in each index condition (see Appendix-2), and demographics. The Hierarchical Model additionally included subgroup membership.
B. *Model Internal Validation.* We used the validation dataset to internally validate the models through the following two measures:
C. Discrimination (model’s ability to distinguish readmitted patients from those not readmitted) was measured using the C-statistic, which is identical to the area under the receiver operating characteristic (ROC) curve. Model discrimination was examined using box plots to show the average risk prediction for patients with and without readmission.
D. Calibration (model’s agreement of the predicted probabilities with the observed risk) was measured using calibration-in-the-large, and calibration slope, which was examined through a calibration plot showing the proportion of patients actually admitted, versus deciles of predicted probability of having readmission. Good calibration is when calibration-in-the-large is close to zero, and the calibration slope is close to one. Since the large sample size overpowered the study, we did not measure the calibration based on statistical significance (e.g., *P* values of the Hosmer-Lemeshow and calibration indices).
E. *Model Comparisons.* We used the chi-squared test to compare the C-statistic of the Standard Model to that of the Hierarchical Model. We also measured the C-statistic of the Standard Model applied to each subgroup separately. This enabled examination of how the Standard Model performed on patient subgroups to identify, for example, which subgroups underperformed when using the current Standard Model.

Because the above models used the feature selection step to select comorbidities for use as independent variables, they differed from those used in the published CMS models. Therefore, to perform a head-to-head comparison with the published CMS models, we additionally developed a logistic regression model using independent variables that were identical to the published CMS model (CMS Standard Model), which was compared to the same model that included subgroup membership (CMS Hierarchical Model). We used the chi-squared test to compare the C-statistic of the CMS Standard Model to that from the CMS Hierarchical Model, in addition to the following measures of model accuracy:

1. Net Reclassification Improvement (NRI) measured the proportion of patients whose predicted probability of readmission improved with reference to actual readmission status. We used two NRI statistics: (a) categorical NRI, which predicted readmission probabilities divided into 10 sequential categories ranging from 0-1, with improvement requiring a shift between categories; and (b) continuous NRI which is based on the proportions of patients with any improved predicted probability of readmission, regardless of the size of that improvement.
2. Integrated Discrimination Improvement (IDI) measured the difference in the average improvement in predicted risks between the CMS Standard Model and the CMS Hierarchical Model.

## APPENDIX-2 Patient Inclusion and Exclusion Criteria

### ICD-9 Codes for Selecting Index Conditions

1. **Hospitalization for COPD** was defined as (1) hospitalization with a primary ICD-9 code for COPD (491.21, 491.22, 491.8, 491.9, 492.8, 493.20, 493.21, 493.22 and 496), or (2) primary ICD-9 codes of 518.81, 518.82, 518.84 or 799.1 and secondary ICD-9 codes for COPD (491.21, 491.22, 493.21 and 493.22).
2. **Hospitalization for CHF** was defined as hospitalization with a primary ICD-9 code for CHF (402.01, 402.01, 402.91, 404.01, 404.03, 404.11, 404.13, 404.91, 404.93 or 428.xx)
3. **Hospitalization for THA/TKA** was defined as hospitalization with a primary ICD-9 code for THA/TKA (81.51 and 81.54). We included admissions only for elective total hip/knee arthroplasty, and those with non-elective were excluded. These included admissions with a diagnosis of a femur, hip, or pelvic fracture, those who received partial hip arthroplasty, revision or resurfacing procedures concurrently with hip/knee arthroplasty, those with malignant bone neoplasm, or with a procedure code for removal of implanted devices/prostheses.

### Inclusion and Exclusion Criteria

For each index condition, we used the same inclusion and exclusion criteria used to develop the CMS models, but with the most recent years (2013-2014) provided by Medicare when we started the project. We used 100% of the 30-day readmitted patients in 2013 and 2014 Medicare claims data, from which we extracted all patients that were admitted to an acute care hospital on or after July 2013-August 2014 with a principal diagnosis of the index condition, were 66 years of age or older, and were enrolled in both Medicare parts A and B fee-for-service plans in the 6 months before admission. Furthermore, we excluded patients who were transferred from other facilities, died during the hospitalization, or transferred to another acute care hospital. Similar to the CMS models, we selected the first admission for patients with multiple admissions during the study period, and did not use Medicare Part D (related to prescription medications).

Next, we extracted 100% controls who were not readmitted for at least 90 days since discharge. CMS uses this 90-day window of no re-admittance to ensure that the controls are substantially free of complications that result in readmission during this period [12, 13]. A small percentage (0.8%) of Medicare patients had “unknown race” for the Race attribute, so we grouped “unknown race” and “other race” and ensured that there was an equal number of them in the cases and control datasets. The low rate of missing data on race had too low a risk for bias to warrant a sensitivity analysis. The following flow charts describe the inclusion and exclusion criteria used to extract cases and controls for COPD, CHF, and THA/TKA, and the respective numbers of patients extracted at each step.

### Patient Inclusion and Exclusion Criteria for COPD Training and Replication Datasets

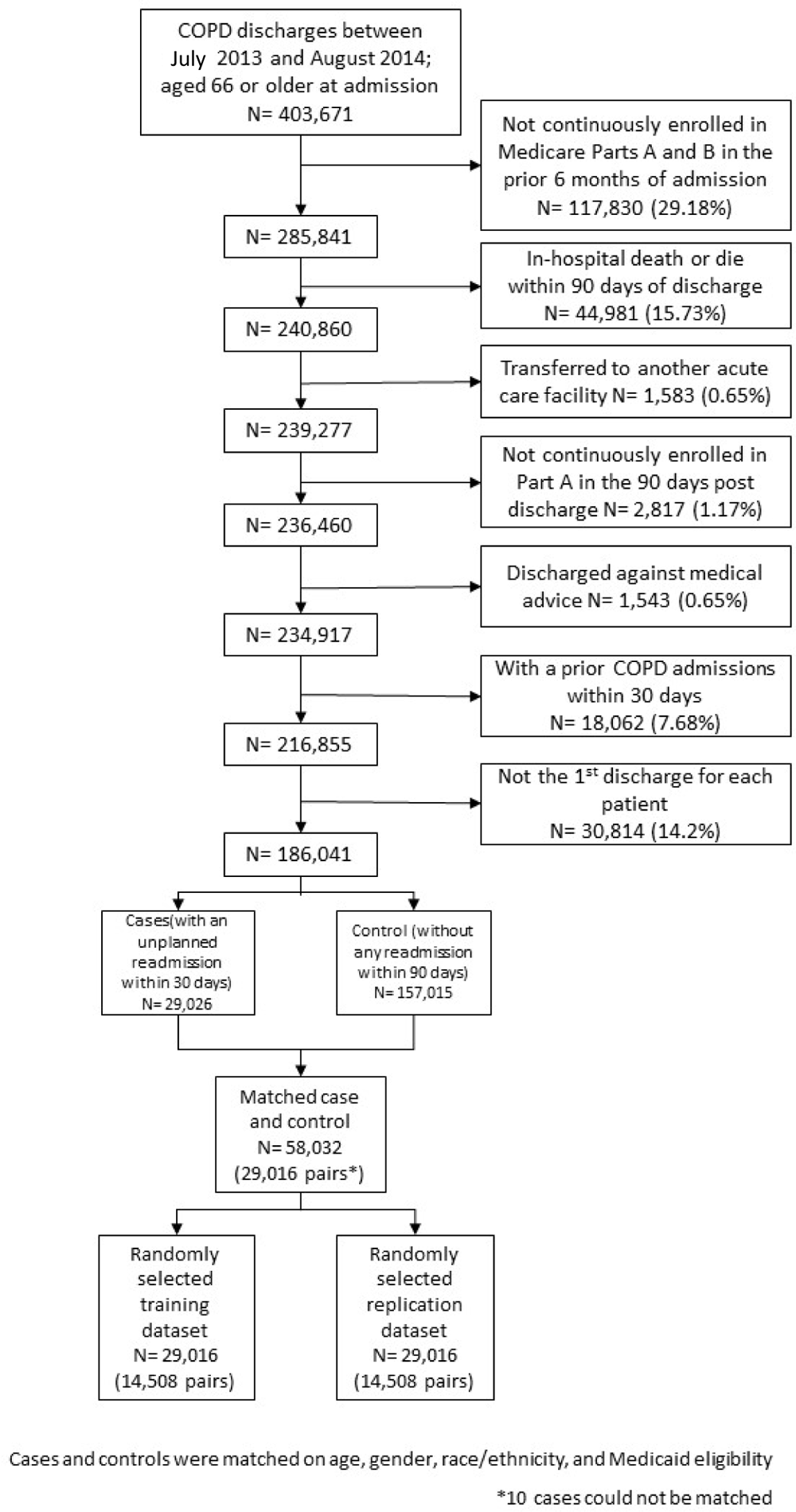

### Patient Inclusion and Exclusion Criteria for CHF Training and Replication Datasets

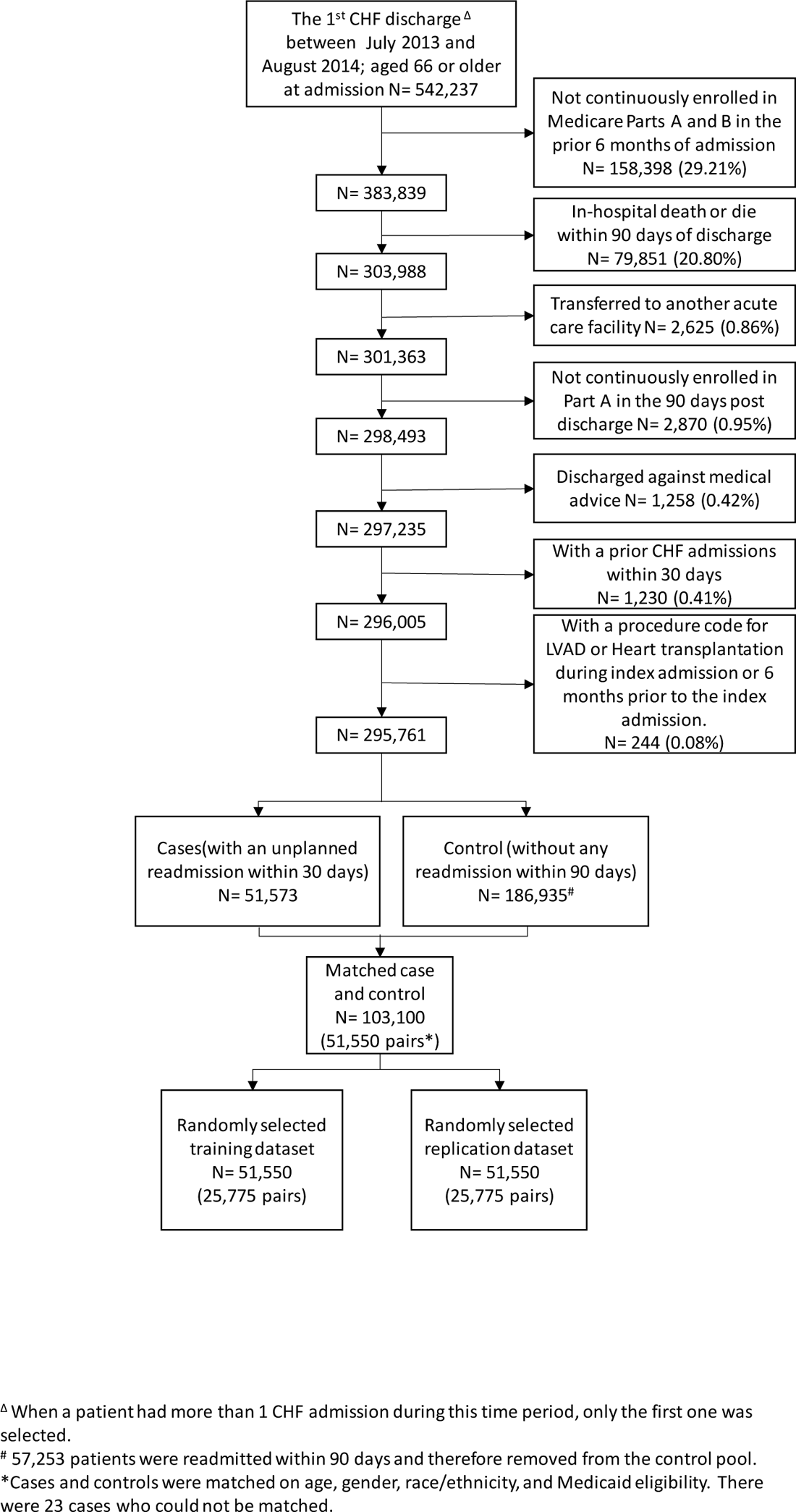

### Patient Inclusion and Exclusion Criteria for THA/TKA Training and Replication Datasets

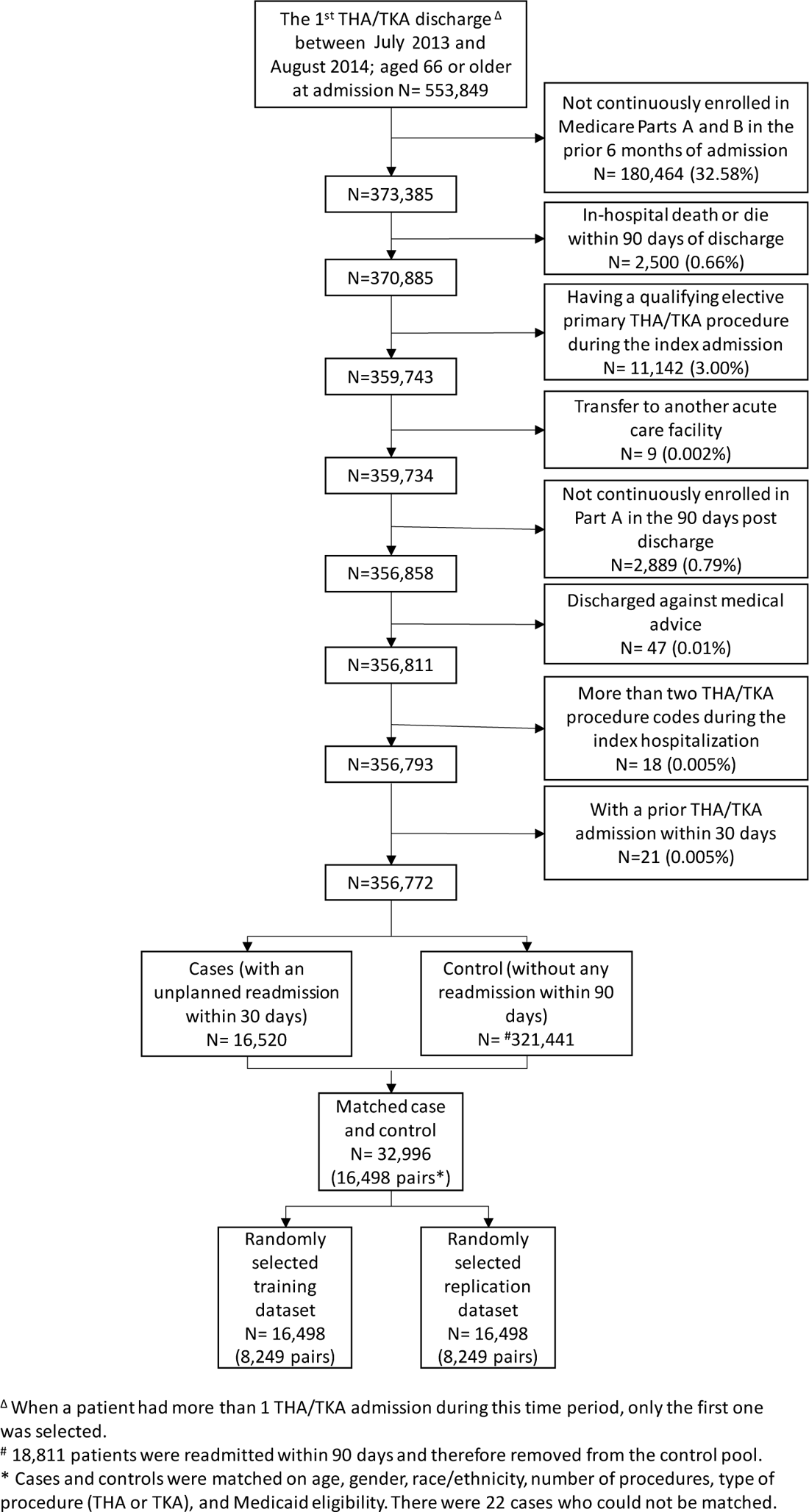

## APPENDIX-3 Variable and Feature Selection

### COPD

The initial set of comorbidities included 45 comorbidities generated from a union of the three comorbidity indices, plus 2 condition-specific comorbidities recommended by the clinicians, resulting in 47 comorbidities. The following feature-selection steps resulted in 30 comorbidities surviving, that were used for the modeling:

1. Removed comorbidities with prevalence less than 1%, resulting in the following that were excluded, leaving 44 comorbidities:
2. Measured the OR of each comorbidity for readmission and excluded the following that were not significant (at the .05 level corrected for multiple testing with Bonferroni), leaving 40 comorbidities:
3. Conducted a two-way co-occurrence test resulting in none being excluded.
4. Conducted a two-way directionality test resulting in the following that were excluded:
5. Repeated steps 2-4 in the replication dataset resulting in 30 comorbidities shown below:

| Excluded Comorbidities | Label |
| --- | --- |
| 1. RespDepend | Respirator dependence/respiratory failure (V22 CC 82-83) |
| 2. Pancreatitis | Chronic pancreatitis (V22 CC 34) |
| 3. HIV_AIDS | HIV/AIDS |

| Excluded Comorbidities | Label |
| --- | --- |
| 1. Neoplasm_other | Other digestive and urinary neoplasms (V22 CC 14) |
| 2. Lymphatic | Lymphatic, head and neck, brain, and other major cancers; breast, colorectal and other cancers and tumors; other respiratory and heart neoplasms |
| 3. Diabetes_wo_comp | Diabetes without complications |
| 4. Rheumatic | Connective Tissue Disease-Rheumatic Disease |

3. Conducted a two-way co-occurrence test resulting in none being excluded. 4. Conducted a two-way directionality test resulting in the following that were excluded:
| Excluded Comorbidities | Label |
| --- | --- |
| 1. Anxiety | Anxiety disorders (V22 CC 62) |
| 2. Brain_disorder | Dementia or other specified brain disorders |
| 3. Liver_disease | Liver disease |
| 4. LungCa | Lung and other severe cancers (V22 CC 9) |
| 5. Metastatic_cancer | Metastatic cancer or acute leukemia |
| 6. Vertebral_fract | Vertebral fractures without spinal cord injury (V22 CC 169) |

5. Repeated steps 2-4 in the replication dataset resulting in 30 comorbidities shown below:
| Final Comorbidities | Label |
| --- | --- |
| 1. Anemias | Iron deficiency or other/unspecified anemias and blood disease |
| 2. Apnea | Sleep apnea |
| 3. Arrhythmias | Specified arrhythmias and other heart rhythm disorders |
| 4. CardioRespShock | Cardio-respiratory failure and shock (V22 CC 84) |
| 5. Cellulitis | Cellulitis, local skin infection (V22 CC 164) |
| 6. Coronary_angina | Coronary atherosclerosis or angina (V22 CC 88-89) |
| 7. Coronary_syndrome | Acute coronary syndrome |
| 8. Depression | Depression (V22 CC 61) |
| 9. Diabetes_w_comp | Diabetes with complications |
| 10. Endocrine_disorder | Endocrine and metabolic disorders; disorders of fluid/electrolyte/acidbase balance |
| 11. GI_other | Other gastrointestinal disorders (V22 CC 38) |
| 12. HD_other | Other and unspecified heart disease (V22 CC 98) |
| 13. Heart_failure | Congestive heart failure |
| 14. Hemiplegia | Hemiplegia, paraplegia, paralysis, functional disability |
| 15. Hypertension_comp | Hypertension complicated |
| 16. Hypertension_Uncomp | Hypertension uncomplicated |
| 17. Infection | Severe infection; other infectious diseases (V22 CC 3-7) |
| 18. Malnutrition | Protein-calorie malnutrition / weight loss |
| 19. Morbid_OB | Morbid obesity; other endocrine/metabolic/nutritional disorders |
| 20. MV | History of mechanical ventilation |
| 21. Neurological_Disorders | Other Neurological Disorders |
| 22. Neuropathy | Polyneuropathy / other neuropathies (V22 CC 75,81) |
| 23. Peptic_ulcer | Peptic ulcer, hemorrhage, other specified gastrointestinal disorders |
| 24. Pneu | Pneumonia (V22 CC 114-116) |
| 25. psych_other | Other psychiatric disorders (V22 CC 63) |
| 26. Psychosis | Drug/alcohol psychosis or dependence |
| 27. Renal_failure | Renal failure |
| 28. Ulcer | Decubitus ulcer or chronic skin ulcer (V22 CC 157-161) |
| 29. Valvular_Disease | Valvular Disease |
| 30. Vascular | Vascular or circulatory disease |

### CHF

The initial set of comorbidities included 42 comorbidities generated from a union of the three comorbidity indices, plus 1 condition-specific comorbidities recommended by the clinicians, resulting in 43 comorbidities. The following feature-selection steps resulted in 37 comorbidities, that were used for the modeling:

1. Removed comorbidities with prevalence less than 1%, resulting in the following that were excluded, leaving 42 comorbidities:
2. Measured the OR of each comorbidity for readmission and excluded the following that were not significant (at the .05 level corrected for multiple testing with Bonferroni), leaving 40 comorbidities that had significant associations with readmission:
3. Conducted a two-way co-occurrence test resulting in none being excluded.
4. Conducted a two-way directionality test, resulting in the following that were excluded leaving 39 variables that were involved in one or more significant direction tests:
5. Repeated steps 2-4 in the replication dataset resulting in 37 comorbidities shown below:

| Excluded Comorbidities | Label |
| --- | --- |
| 1. HIV_AIDS | HIV/AIDS |

| Excluded Comorbidities | Label |
| --- | --- |
| 1. Metastatic_cancer | Metastatic cancer or acute leukemia |
| 2. Diabetes_wo_comp | Diabetes Mellitus without Complication |

| Excluded Comorbidities | Label |
| --- | --- |
| 1. Rheumatic | Rheumatic Disease |

5. Repeated steps 2-4 in the replication dataset resulting in 37 comorbidities shown below:
| Final Comorbidities | Label |
| --- | --- |
| 1. Coronary_angina | Coronary atherosclerosis or angina (CC 88-89) |
| 2. CABG | History of coronary artery bypass graft (CABG) surgery |
| 3. COPD | Chronic Obstructive Pulmonary Disease (CC 111) |
| 4. CardioRespShock | Cardio-respiratory failure and shock |
| 5. Depression | Depression (CC 61) |
| 6. Dialysis | Dialysis status (CC 134) |
| 7. GI_other | Other gastrointestinal disorders (CC 38) |
| 8. Hypertension_Comp | Hypertension Complicated |
| 9. Hypertension_Uncomp | Hypertension Uncomplicated |
| 10. Hypothyroidism | Hypothyroidism |
| 11. Nephritis | Nephritis (CC 141) |
| 12. Obesity | Obesity |
| 13. Neuro_disorders | Other Neurological Disorders |
| 14. HD_other | Other and unspecified heart disease (CC 98) |
| 15. Psych_other | Other psychiatric disorders (CC 63) |
| 16. Urinary_tract_disorder | Other urinary tract disorders (CC 145) |
| 17. Pneu | Pneumonia (CC 114-116) |
| 18. Renal_failure | Renal failure (CC 135-140) |
| 19. Ulcer | Decubitus ulcer or chronic skin ulcer (CC 157-161) |
| 20. Coronary_syndrome | Acute coronary syndrome |
| 21. Psychosis | Drug/alcohol abuse/dependence/psychosis |
| 22. Anemias | Iron deficiency or other/unspecified anemias and blood disease |
| 23. Arrhythmia | Specified arrhythmias and other heart rhythm disorders |
| 24. CHF | Congestive heart failure |
| 25. Cancer | Cancer |
| 26. Stroke | Stroke |
| 27. Brain_disorders | Dementia or other specified brain disorders |
| 28. Diabetes_w_comp | Diabetes Mellitus Complicated |
| 29. Endocrine_disorders | Other significant endocrine and metabolic disorders; disorders of fluid/electrolyte/acid base balance |
| 30. Liver_disease | Liver or biliary disease |
| 31. Malnutrition | Protein-calorie malnutrition |
| 32. Hemiplegia | Hemiplegia, paraplegia, paralysis, functional disability |
| 33. Peptic_ulcer | Peptic ulcer, hemorrhage, other specified gastrointestinal disorders |
| 34. Vascular | Vascular or circulatory disease |
| 35. Psychiatric_disorders | Major psychiatric disorders |
| 36. Hematological | Severe hematological disorders |
| 37. Valvular_disease | Valvular and Rheumatic Heart Disease |

### TKA/THA

The initial set of comorbidities included 39 comorbidities generated from a union of the three comorbidity indices, plus 2 condition-specific comorbidities recommended by the clinicians, resulting in 41 comorbidities. The following feature-selection steps resulted in 11 comorbidities, that were used for the modeling:

1. Removed comorbidities with prevalence less than 1%, resulting in the following that were excluded, leaving 30 comorbidities:
2. Measured the OR of each comorbidity for readmission (at the .05 level corrected for multiple testing with Bonferroni), leaving all 30 comorbidities that had significant associations with readmission
3. Conducted a two-way co-occurrence test resulting in none being excluded.
4. Conducted a two-way directionality test, resulting in the following that were excluded leaving 19 variables that were involved in one or more significant direction tests:
5. Repeated steps 2 through 4 in the replicate dataset resulting in 11 comorbidities shown below:

| Excluded Comorbidities | Label |
| --- | --- |
| 1. Other_Hip_Cong_Def | Other congenital deformity of hip (joint) |
| 2. Post_Trau_Osteoarthritis | Post traumatic osteoarthritis |
| 3. Dialysis_status | Dialysis status (CC 134) |
| 4. Blood_Loss_Anemia | Blood Loss Anemia |
| 5. Alcohol_Abuse | Alcohol Abuse |
| 6. Drug_Abuse | Drug Abuse |
| 7. HIV_AIDS | HIV/AIDS |
| 8. Metastatic_cancer | Metastatic cancer or acute leukemia |
| 9. Hemiplegia | Hemiplegia, paraplegia, paralysis, functional disability |
| 10. Liver_disease | Liver disease |
| 11. Peptic_ulcer | Peptic Ulcer Disease |

| Excluded Comorbidities | Label |
| --- | --- |
| 1. Brain_disorders | Dementia or other specified brain disorders |
| 2. Cancer | Cancer |
| 3. Cellulitis | Cellulitis, local skin infection (CC 164) |
| 4. Deficiency_Anemia | Deficiency Anemia |
| 5. Diab_wo_comp | Diabetes mellitus |
| 6. Hematological | Severe hematological disorders |
| 7. Malnutrition | Protein-calorie malnutrition |
| 8. Neuro_disorders | Other Neurological Disorders |
| 9. Stroke | Stroke |
| 10. Ulcer | Decubitus ulcer or chronic skin ulcer (CC 157-161) |
| 11. Valvular_disease | Valvular Disease |

5. Repeated steps 2 through 4 in the replicate dataset resulting in 11 comorbidities shown below:
| Final Comorbidities | Label |
| --- | --- |
| 1. Arrhythmia | Specified arrhythmias and other heart rhythm disorders |
| 2. CHF | Congestive heart failure |
| 3. COPD | Chronic Obstructive Pulmonary Disease |
| 4. Coronary_angina | Coronary atherosclerosis or angina |
| 5. Endocrine_disorders | Other significant endocrine and metabolic disorders; disorders of fluid/electrolyte/acidbase balance |
| 6. Hypertension_Comp | Hypertension complicated |
| 7. Hypertension_Uncomp | Hypertension Uncomplicated |
| 8. Major_Symp_Abnormalities | Major symptoms, abnormalities (CC 178) |
| 9. Morbid_OB | Morbidity obesity |
| 10. Psychiatric_disorders | Major psychiatric disorders |
| 11. Renal_failure | Renal failure (CC 135-140) |

## APPENDIX-4 Classification Modeling

### Multinomial Logistic Regression Coefficients

#### COPD

|  | **Bicluster-2** | **Bicluster-3** | **Bicluster-4** |
| --- | --- | --- | --- |
| (Intercept) | 9.478442033 | -15.60229477 | -20.58911845 |
| MV | 15.37003888 | 31.79526753 | 174.7625575 |
| Apnea | 14.73866236 | 30.13129952 | 174.8192981 |
| Infection | 13.07141525 | 30.94167533 | 175.7976241 |
| GI_other | -138.0330506 | -122.1606071 | -141.8251611 |
| Depression | 11.44920157 | 194.57748 | 15.50399067 |
| Psych_other | 6.633772193 | 193.1120262 | 15.01395622 |
| Neuropathy | 4.22511563 | 191.7033722 | 11.82453364 |
| CardioRespShock | 15.935738 | 30.78720915 | 177.4810561 |
| Coronary_angina | -140.3504037 | -121.6295958 | -139.1018879 |
| HD_other | 12.65500876 | 22.79910161 | 173.1189166 |
| Pneu | 12.40491598 | 192.8944261 | 14.40776138 |
| Ulcer | 25.40389722 | 36.65033091 | 174.4945337 |
| Cellulitis | 10.19338572 | 25.35264195 | 174.1779677 |
| Valvular_Disease | 179.470279 | 29.79933499 | 12.07687072 |
| Hypertension_Uncomp | -144.0822947 | -135.252917 | -149.1763872 |
| Hypertension_comp | 169.9103199 | 9.909139715 | -15.01176101 |
| Neurological_Disorders | 11.98793007 | 27.3962048 | 174.152075 |
| Diabetes_w_comp | 179.9458627 | 32.79407674 | 10.94569788 |
| Malnutrition | 2.619696434 | 190.2553747 | 10.57634814 |
| Morbid_OB | -144.7853066 | -126.4723011 | -150.75158 |
| Endocrine_disorder | 18.25087715 | 30.17692505 | 173.8734803 |
| Peptic_ulcer | 14.57401571 | 33.3138924 | 174.6959617 |
| Anemias | 180.37182 | 32.01649034 | 11.85268168 |
| Psychosis | 0.808153461 | 190.136092 | 8.470673123 |
| Hemiplegia | 22.05837142 | 36.91425527 | 173.7515174 |
| Heart_failure | 179.4801884 | 29.61286201 | 6.090102717 |
| Coronary_syndrome | 17.65746601 | 31.18056088 | 177.0738381 |
| Arrhythmias | -140.5676174 | -122.0347651 | -139.1832525 |
| Vascular | 12.82066036 | 29.47640378 | 174.8331118 |
| Renal_failure | 181.4635165 | 35.07327525 | 9.594086268 |

#### CHF

|  | **Bicluster-2** | **Bicluster-3** | **Bicluster-4** |
| --- | --- | --- | --- |
| (Intercept) | 4.228411649 | 3.221587055 | 4.404809652 |
| CABG | -16.88332455 | -16.46705046 | -16.90410559 |
| GI_other | 1.608383181 | 18.42437593 | 2.537152385 |
| Depression | 1.744237257 | 18.57833534 | 2.132954431 |
| Psych_other | 1.565344584 | 18.89075678 | 1.851558983 |
| CardioRespShock | 1.899086259 | 2.614460813 | 20.82096349 |
| Coronary_angina | -16.52943558 | -16.50069914 | -16.28824682 |
| HD_other | 1.630347764 | 1.900983651 | 20.69135601 |
| COPD | 2.208517874 | 18.8515632 | 2.812111665 |
| Pneu | 1.368125374 | 1.230710494 | 20.35722646 |
| Dialysis_status | 21.64550697 | 7.573268069 | 6.356824836 |
| Renal_failure | 16.64635654 | 3.207105993 | 2.875810167 |
| Nephritis | 24.05426922 | 11.91409993 | 8.430710694 |
| Other_uri_tract_disorders | 17.28359996 | 3.468138309 | 2.68316497 |
| Ulcer | 18.62035155 | 4.612512833 | 4.621717632 |
| Hypertension_Uncomp | -16.17798732 | -17.43341906 | -17.17899564 |
| Hypertension_Comp | 18.21349274 | 2.666169079 | 2.719974312 |
| Neuro_Disorders | 3.174148638 | 3.426243554 | 21.7581539 |
| Hypothyroidism | 1.532038616 | 18.6465425 | 2.742755582 |
| Obesity | 16.77179346 | 3.477665718 | 1.851913596 |
| Cancer | 1.825845108 | 18.78533294 | 2.280566193 |
| Diabetes_w_comp | 16.97604199 | 3.131645807 | 2.25066864 |
| Malnutrition | 1.654276729 | 2.26075743 | 20.16266194 |
| Endocrine_disorder | 16.95913583 | 3.414523477 | 2.762293049 |
| Liver_disease | 0.509273487 | 1.346254075 | 19.37619302 |
| Peptic_ulcer | 1.882489155 | 2.868064904 | 20.99453939 |
| Hematological | 1.215479083 | 1.623296079 | 20.18186306 |
| Anemias | 16.84448889 | 2.650113464 | 2.132489404 |
| Brain_disorders | 1.214570676 | 19.0044895 | 2.119832832 |
| Psychosis | 1.612149611 | 18.59666356 | 2.038965354 |
| Psychiatric_disorders | 3.533134983 | 19.8653935 | 3.164125739 |
| Hemiplegia | 2.634196819 | 3.196095963 | 21.40555332 |
| CHF | -16.83591928 | -17.14949663 | -16.56386079 |
| Coronary_syndrome | 1.423033472 | 2.813198347 | 20.7510375 |
| Valvular_disease | -16.84402957 | -16.55559304 | -16.67291059 |
| Arrhythmia | -17.41765184 | -17.22878444 | -17.44203684 |
| Stroke | 1.248759632 | 1.486455436 | 19.78749343 |
| Vascular | 0.908845182 | 1.356092137 | 19.93213936 |

#### TKA/THA

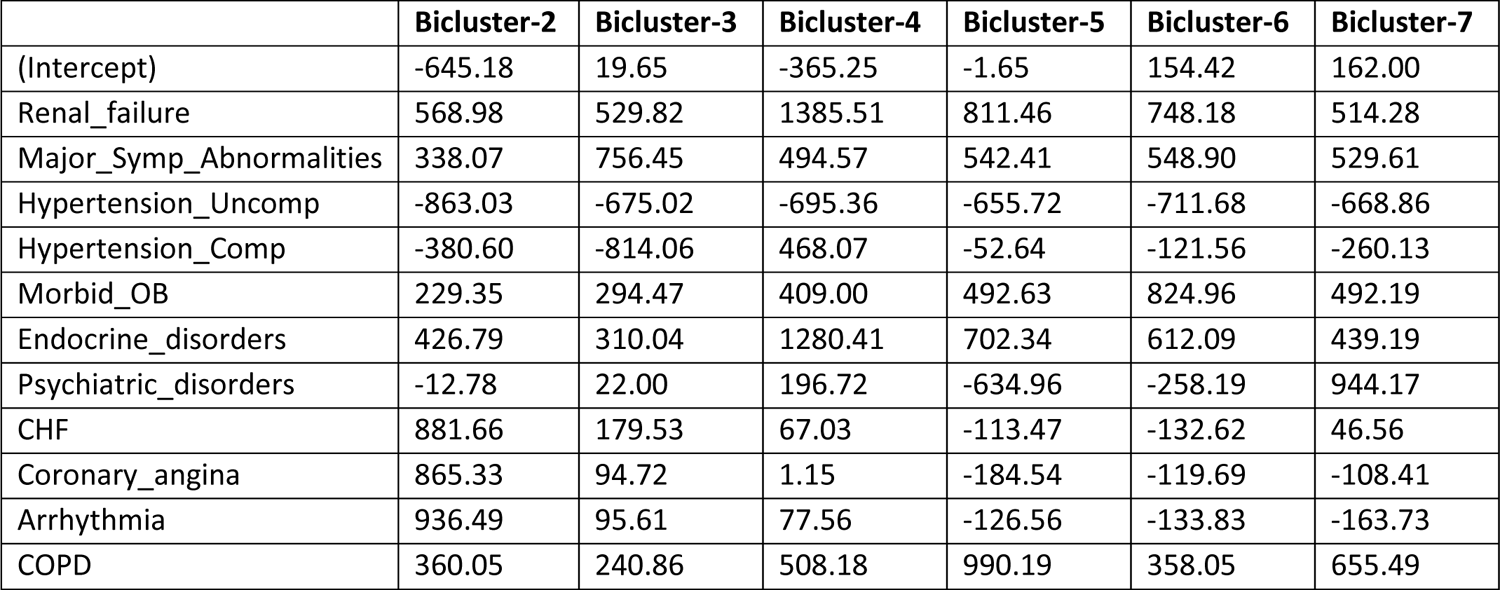

|  | Bicluster-2 | Bicluster-3 | Bicluster-4 | Bicluster-5 | Bicluster-6 | Bicluster-7 |
| --- | --- | --- | --- | --- | --- | --- |
| (Intercept) | -645.18 | 19.65 | -365.25 | -1.65 | 154.42 | 162.00 |
| Renal_failure | 568.98 | 529.82 | 1385.51 | 811.46 | 748.18 | 514.28 |
| Major_Symp_Abnormalities | 338.07 | 756.45 | 494.57 | 542.41 | 548.90 | 529.61 |
| Hypertension_Uncomp | -863.03 | -675.02 | -695.36 | -655.72 | -711.68 | -668.86 |
| Hypertension_Comp | -380.60 | -814.06 | 468.07 | -52.64 | -121.56 | -260.13 |
| Morbid_OB | 229.35 | 294.47 | 409.00 | 492.63 | 824.96 | 492.19 |
| Endocrine_disorders | 426.79 | 310.04 | 1280.41 | 702.34 | 612.09 | 439.19 |
| Psychiatric_disorders | -12.78 | 22.00 | 196.72 | -634.96 | -258.19 | 944.17 |
| CHF | 881.66 | 179.53 | 67.03 | -113.47 | -132.62 | 46.56 |
| Coronary_angina | 865.33 | 94.72 | 1.15 | -184.54 | -119.69 | -108.41 |
| Arrhythmia | 936.49 | 95.61 | 77.56 | -126.56 | -133.83 | -163.73 |
| COPD | 360.05 | 240.86 | 508.18 | 990.19 | 358.05 | 655.49 |

### Subgroup Risk

#### COPD

The COPD dataset included 186041 total patients, of which 29026 were cases (15.6%). The following are the percentage of cases in each bicluster (subgroup risk) after classification of the 100% cases and 100% controls by the classification model, and then juxtaposed with the visualization of the respective patient subgroups:

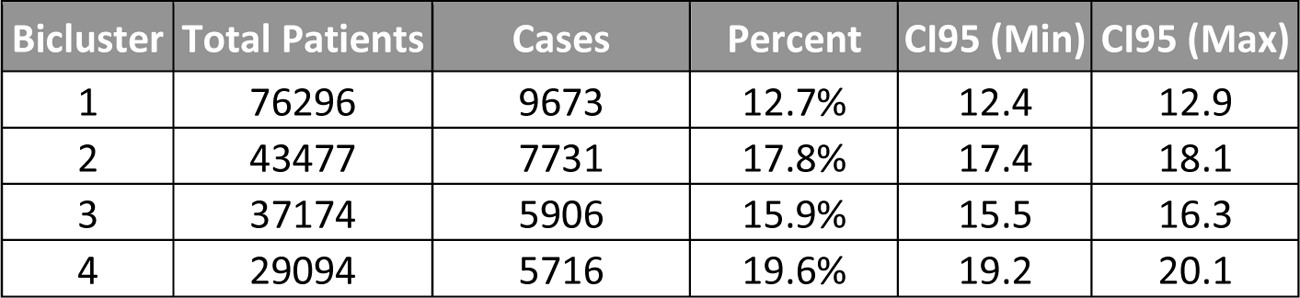

| Bicluster | Total Patients | Cases | Percent | CI95 (Min) | CI95 (Max) |
| --- | --- | --- | --- | --- | --- |
| 1 | 76296 | 9673 | 12.7% | 12.4 | 12.9 |
| 2 | 43477 | 7731 | 17.8% | 17.4 | 18.1 |
| 3 | 37174 | 5906 | 15.9% | 15.5 | 16.3 |
| 4 | 29094 | 5716 | 19.6% | 19.2 | 20.1 |

#### CHF

The CHF dataset included 295761 total patients, of which 51573 were cases (17.4%). The following are the percentage of cases in each bicluster (subgroup risk) after classification of the 100% cases and 100% controls into subgroups by the classification model, and juxtaposed with the visualization of the respective patient subgroups:

| Bicluster | Total Patients | Cases | Percent | CI95 (Min) | CI95 (Max) |
| --- | --- | --- | --- | --- | --- |
| 1 | 9673 | 76296 | 12.7% | 12.4 | 12.9 |
| 2 | 7731 | 43477 | 17.8% | 17.4 | 18.1 |
| 3 | 5906 | 37174 | 15.9% | 15.5 | 16.3 |
| 4 | 5716 | 29094 | 19.6% | 19.2 | 20.1 |

#### THA/TKA

The TKA/THA dataset included 356772 total patients, of which 16520 were cases (4.6%). The following are the percentage of cases in each bicluster (subgroup risk) after classification of 100% cases and 100% controls by the classification model, and then juxtaposed with the visualization of the respective patient subgroups:

| Bicluster | Total Patients | Cases | Percent | CI95 (Min) | CI95 (Max) |
| --- | --- | --- | --- | --- | --- |
| 1 | 2157 | 57045 | 3.8% | 3.6 | 3.9 |
| 2 | 2317 | 33535 | 6.9% | 6.6 | 7.2 |
| 3 | 2255 | 55260 | 4.1% | 3.9 | 4.2 |
| 4 | 2135 | 29475 | 7.2% | 7 | 7.5 |
| 5 | 2003 | 30856 | 6.5% | 6.2 | 6.8 |
| 6 | 1945 | 43185 | 4.5% | 4.3 | 4.7 |
| 7 | 3708 | 107416 | 3.5% | 3.3 | 3.6 |

## APPENDIX-5 Predictive Modeling

### COPD

#### Discrimination

The following are box plots of risk prediction by readmission status for the Standard Model and the Hierarchical Model.

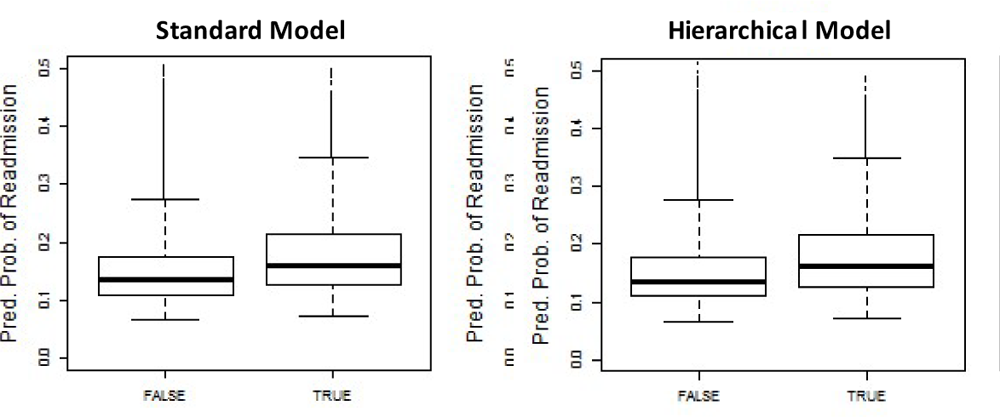

#### Calibration

The following are calibration plots showing logistic regressions relating readmission status to the logistic quantile of the predicted probability of readmission yield regression lines with specified intercept and slope (the ideal regression line would have intercept=0 and slope=1, which is shown shaded for reference). The histograms at the bottom of each graph reflect the frequency of modeled data, horizontal axes show predicted probability, vertical axes show actual probability, and the axes of all figures are constrained to range from 0 to .5.

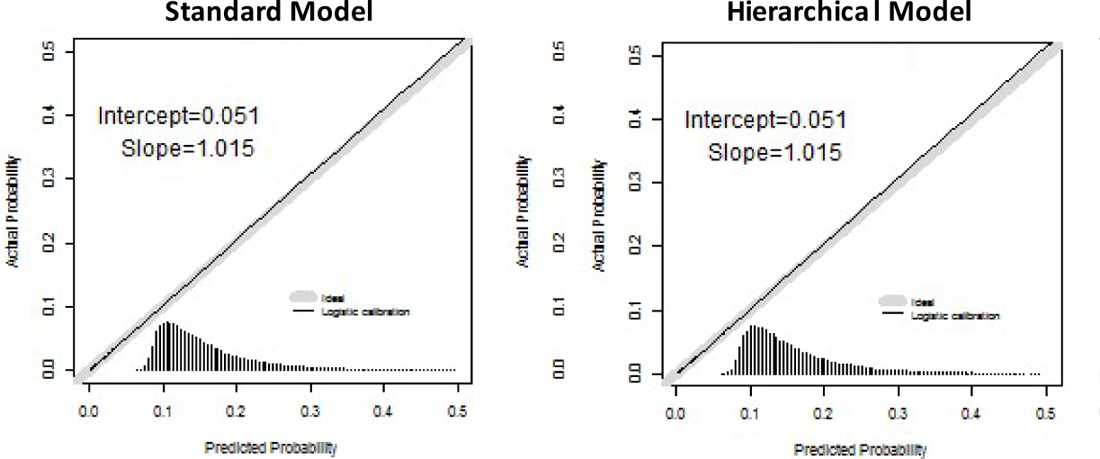

#### Coefficients

Logistic regression coefficients relating readmission status to the logistic quantile of the predicted probability of readmission for each model, with standard errors.

|  | Intercept (Standard Error) | Slope (Standard Error) |
| --- | --- | --- |
| Standard Model | 0.051 (0.049) | 1.015 (0.029) |
| Hierarchical Model | 0.051 (0.049) | 1.015 (0.029) |

### Model Coefficients for Standard Model (COPD)

|  |  |
| --- | --- |
| (Intercept) | -2.136379215 |
| sex2 | -0.018974047 |
| AgeAdm | -0.006994626 |
| RACE51 | 0.295066886 |
| RACE52 | 0.341204445 |
| RACE53 | 0.171206943 |
| RACE55 | 0.273604446 |
| MV | 0.117480441 |
| Apnea | -0.012262741 |
| Infection | 0.026450984 |
| GI_other | 0.087222647 |
| Depression | 0.080231389 |
| Psych_other | 0.082623979 |
| Neuropathy | 0.069716803 |
| CardioRespShock | 0.228794257 |
| Coronary_angina | 0.048182744 |
| HD_other | 0.015511356 |
| Pneu | 0.057607182 |
| Ulcer | 0.091465998 |
| Cellulitis | 0.095049297 |
| Valvular_Disease | 0.078134984 |
| Hypertension_Uncomp | 0.087684427 |
| Hypertension_comp | 0.143660314 |
| Neurological_Disorders | 0.075023091 |
| Diabetes_w_comp | 0.077946469 |
| Malnutrition | 0.095555424 |
| Morbid_OB | -0.056028398 |
| Endocrine_disorder | 0.137464197 |
| Peptic_ulcer | 0.114498859 |
| Anemias | 0.1663521 |
| Psychosis | 0.13049134 |
| Hemiplegia | 0.113401107 |
| Heart_failure | 0.23205475 |
| Coronary_syndrome | 0.110911596 |
| Arrhythmias | 0.188301605 |
| Vascular | 0.090867219 |
| Renal_failure | 0.11154648 |

### Model Coefficients for Hierarchical Model (COPD)

|  |  |
| --- | --- |
| (Intercept) | -2.170381336 |
| sex2 | -0.018743606 |
| AgeAdm | -0.00702047 |
| RACE51 | 0.29629354 |
| RACE52 | 0.342336002 |
| RACE53 | 0.172316947 |
| RACE55 | 0.275083731 |
| MV | 0.112639359 |
| Apnea | -0.017323465 |
| Infection | 0.02133693 |
| GI_other | 0.095383617 |
| Depression | 0.072418589 |
| Psych_other | 0.075096553 |
| Neuropathy | 0.063679331 |
| CardioRespShock | 0.224665848 |
| Coronary_angina | 0.056139938 |
| HD_other | 0.011352801 |
| Pneu | 0.052053617 |
| Ulcer | 0.089352719 |
| Cellulitis | 0.091640641 |
| Valvular_Disease | 0.074608627 |
| Hypertension_Uncomp | 0.100372891 |
| Hypertension_comp | 0.143513998 |
| Neurological_Disorders | 0.070642457 |
| Diabetes_w_comp | 0.076044197 |
| Malnutrition | 0.089847317 |
| Morbid_OB | -0.045581819 |
| Endocrine_disorder | 0.132138897 |
| Peptic_ulcer | 0.1109747 |
| Anemias | 0.162850779 |
| Psychosis | 0.124238457 |
| Hemiplegia | 0.109511728 |
| Heart_failure | 0.228425309 |
| Coronary_syndrome | 0.106258 |
| Arrhythmias | 0.19629312 |
| Vascular | 0.085350053 |
| Renal_failure | 0.106363156 |
| PredCluster2 | 0.042362651 |
| PredCluster3 | 0.05007554 |
| PredCluster4 | 0.052171757 |

### Standard Model and Hierarchical Model for COPD

The following table shows C-statistics for the Standard Model and the Hierarchical Model.

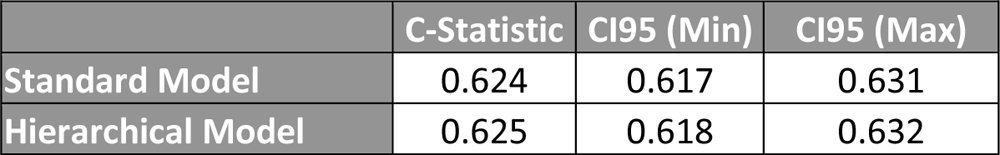

The following table shows C-statistics for the Standard Model used to predict readmission for patients in each bicluster separately.

|  | C-Statistic | CI95 (Min) | CI95 (Max) |
| --- | --- | --- | --- |
| Bicluster 1 | 0.596 | 0.584 | 0.608 |
| Bicluster 2 | 0.610 | 0.596 | 0.624 |
| Bicluster 3 | 0.616 | 0.600 | 0.632 |
| Bicluster 4 | 0.636 | 0.620 | 0.652 |

### CMS Models (CMS Standard Model and CMS Hierarchical Model) for COPD

The following table shows C-statistics for the CMS Standard Model and the CMS Hierarchical Model.

CMS Models (CMS Standard Model and CMS Hierarchical Model) for COPD The following table shows C-statistics for the CMS Standard Model and the CMS Hierarchical Model.
|  | C-Statistic | CI95 (Min) | CI95 (Max) |
| --- | --- | --- | --- |
| CMS Standard Model | 0.622 | 0.615 | 0.629 |
| CMS Hierarchical Model | 0.622 | 0.615 | 0.629 |

The following table shows C-statistics for the CMS Standard Model used to predict readmission for patients in each bicluster separately.

|  | C-Statistic | CI95 (Min) | CI95 (Max) |
| --- | --- | --- | --- |
| Bicluster 1 | 0.587 | 0.575 | 0.599 |
| Bicluster 2 | 0.611 | 0.598 | 0.625 |
| Bicluster 3 | 0.620 | 0.604 | 0.636 |
| Bicluster 4 | 0.638 | 0.622 | 0.654 |

### Model Coefficients for CMS Standard Model (COPD) [14]

|  |  |
| --- | --- |
| (Intercept) | -2.188594326 |
| V1 | -0.004884898 |
| V2 | 0.173933436 |
| V3 | -0.004371274 |
| V4 | 0.014983286 |
| V5 | 0.158622714 |
| V6 | 0.197266795 |
| V7 | 0.082435098 |
| V8 | -0.042473792 |
| V9 | 0.075588823 |
| V10 | 0.089748913 |
| V11 | -0.042615016 |
| V12 | 0.146684496 |
| V13 | 0.381543598 |
| V14 | 0.085139806 |
| V15 | 0.075183842 |
| V16 | 0.114503012 |
| V17 | 0.16450527 |
| V18 | -0.034979967 |
| V19 | 0.198698509 |
| V20 | 0.100948646 |
| V21 | 0.069808662 |
| V22 | 0.023636931 |
| V23 | 0.069163157 |
| V24 | 0.111452974 |
| V25 | 0.064113847 |
| V26 | -0.039814801 |
| V27 | 0.173239512 |
| V28 | 0.212336424 |
| V29 | 0.066160516 |
| V30 | 0.109441845 |
| V31 | 0.190317991 |
| V32 | 0.043561901 |
| V33 | 0.015612425 |
| V34 | 0.074089074 |
| V35 | 0.069126003 |
| V36 | 0.036551409 |
| V37 | 0.174960292 |
| V38 | 0.102815442 |
| V39 | 0.082962429 |
| V40 | 0.127491216 |

### Model Coefficients for CMS Hierarchical Model (COPD)

|  |  |
| --- | --- |
| (Intercept) | -2.222560918 |
| V1 | -0.005037176 |
| V2 | 0.164652122 |
| V3 | -0.011571161 |
| V4 | 0.007278066 |
| V5 | 0.159430966 |
| V6 | 0.197452639 |
| V7 | 0.083353046 |
| V8 | -0.042706545 |
| V9 | 0.071953525 |
| V10 | 0.098535986 |
| V11 | -0.030172663 |
| V12 | 0.143262108 |
| V13 | 0.382813245 |
| V14 | 0.081387202 |
| V15 | 0.086119119 |
| V16 | 0.11252111 |
| V17 | 0.150832777 |
| V18 | -0.035543286 |
| V19 | 0.207400734 |
| V20 | 0.10019542 |
| V21 | 0.07625943 |
| V22 | 0.023819484 |
| V23 | 0.076936963 |
| V24 | 0.105746192 |
| V25 | 0.07104448 |
| V26 | -0.033998175 |
| V27 | 0.168407005 |
| V28 | 0.202506355 |
| V29 | 0.063304577 |
| V30 | 0.116819091 |
| V31 | 0.195860925 |
| V32 | 0.037067697 |
| V33 | 0.014270778 |
| V34 | 0.066636971 |
| V35 | 0.069127715 |
| V36 | 0.043952171 |
| V37 | 0.129843017 |
| V38 | 0.099065392 |
| V39 | 0.078360538 |
| V40 | 0.127042521 |
| PredCluster2 | 0.098236779 |
| PredCluster3 | 0.014085719 |
| PredCluster4 | 0.072649738 |

### CHF

#### Discrimination

The following are box plots of risk prediction by readmission status for the Standard Model and the Hierarchical Model.

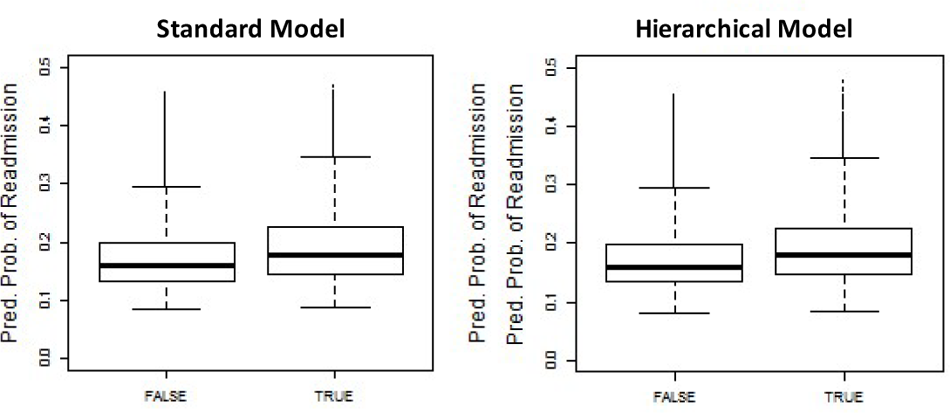

### Calibration

The following are calibration plots showing logistic regressions relating readmission status to the logistic quantile of the predicted probability of readmission yield regression lines with specified intercept and slope (the ideal regression line would have intercept=0 and slope=1, which is shown shaded for reference). The histograms at the bottom of each graph reflect the frequency of modeled data, horizontal axes show predicted probability, vertical axes show actual probability, and the axes of all figures are constrained to range from 0 to .5.

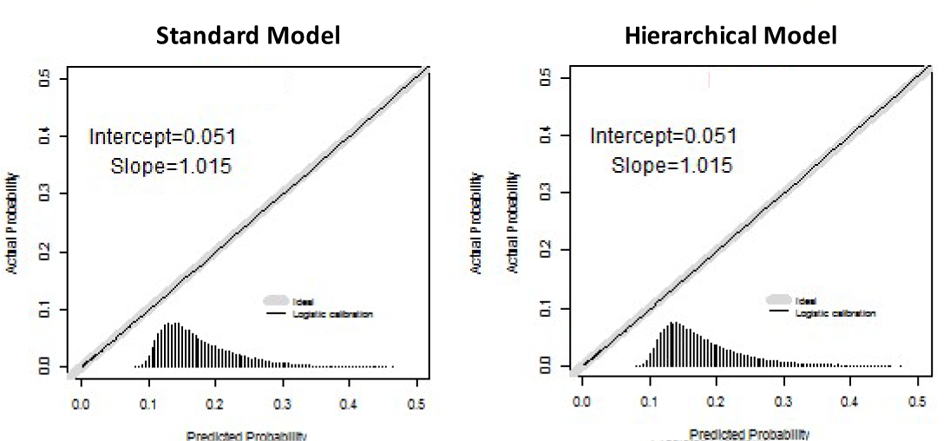

### Coefficients

The following are the logistic regression coefficients relating readmission status to the logistic quantile of the predicted probability of readmission for each model, with standard errors.

|  | Intercept (Standard Error) | Slope (Standard Error) |
| --- | --- | --- |
| Standard Model | .009 (0.042) | 1.013 (0.027) |
| Hierarchical Model | 0.008 (0.042) | 1.013 (0.027) |

### Model Coefficients for Standard Model (CHF)

|  |  |
| --- | --- |
| (Intercept) | -1.475389967 |
| sex2 | 0.074653204 |
| AgeAdm | -0.008514725 |
| RACE51 | -0.036053802 |
| RACE52 | 0.040134152 |
| RACE53 | -0.079761067 |
| RACE55 | 0.072722149 |
| CABG | 0.030894001 |
| GI_other | 0.056464897 |
| Depression | 0.025005144 |
| Psych_other | 0.045337378 |
| CardioRespShock | 0.048277891 |
| Coronary_angina | 0.055087252 |
| HD_other | 0.080761149 |
| COPD | 0.149282237 |
| Pneu | 0.081351108 |
| Dialysis_status | 0.173258929 |
| Renal_failure | 0.140937411 |
| Nephritis | 0.012307007 |
| Other_uri_tract_disorders | 0.065712737 |
| Ulcer | 0.108579462 |
| Hypertension_Uncomp | 0.041031817 |
| Hypertension_Comp | 0.105063607 |
| Neuro_Disorders | 0.032623414 |
| Hypothyroidism | 0.024951326 |
| Obesity | 0.026406255 |
| Cancer | 0.028160953 |
| Diabetes_w_comp | 0.085693489 |
| Malnutrition | 0.038386944 |
| Endocrine_disorder | 0.093422704 |
| Liver_disease | 0.051990801 |
| Peptic_ulcer | 0.053199182 |
| Hematological | 0.045559899 |
| Anemias | 0.123037859 |
| Brain_disorders | -0.00749815 |
| Psychosis | 0.093703137 |
| Psychiatric_disorders | 0.072868486 |
| Hemiplegia | 0.057172104 |
| CHF | 0.106985057 |
| Coronary_syndrome | 0.096953224 |
| Valvular_disease | 0.018311001 |
| Arrhythmia | 0.00786506 |
| Stroke | -0.012731604 |
| Vascular | 0.061193164 |

### Model Coefficients for Hierarchical Model (CHF)

|  |  |
| --- | --- |
| (Intercept) | -1.449848979 |
| sex2 | 0.074706479 |
| AgeAdm | -0.008504158 |
| RACE51 | -0.035610416 |
| RACE52 | 0.04010896 |
| RACE53 | -0.079311376 |
| RACE55 | 0.073284464 |
| CABG | 0.024674808 |
| GI_other | 0.061045731 |
| Depression | 0.029184869 |
| Psych_other | 0.049224598 |
| CardioRespShock | 0.057426429 |
| Coronary_angina | 0.048052865 |
| HD_other | 0.089668573 |
| COPD | 0.154094768 |
| Pneu | 0.089524374 |
| Dialysis_status | 0.164539306 |
| Renal_failure | 0.139877836 |
| Nephritis | 0.004910838 |
| Other_uri_tract_disorders | 0.062840363 |
| Ulcer | 0.105143831 |
| Hypertension_Uncomp | 0.032829812 |
| Hypertension_Comp | 0.100497599 |
| Neuro_Disorders | 0.040022189 |
| Hypothyroidism | 0.029106384 |
| Obesity | 0.022538905 |
| Cancer | 0.031985591 |
| Diabetes_w_comp | 0.082993693 |
| Malnutrition | 0.046167614 |
| Endocrine_disorder | 0.091753086 |
| Liver_disease | 0.060002993 |
| Peptic_ulcer | 0.061215338 |
| Hematological | 0.054764671 |
| Anemias | 0.12180882 |
| Brain_disorders | -0.003163932 |
| Psychosis | 0.097984879 |
| Psychiatric_disorders | 0.075834723 |
| Hemiplegia | 0.065565413 |
| CHF | 0.10067029 |
| Coronary_syndrome | 0.107487626 |
| Valvular_disease | 0.012129124 |
| Arrhythmia | 0.001138884 |
| Stroke | -0.003992176 |
| Vascular | 0.069322294 |
| PredCluster2 | -0.013499297 |
| PredCluster3 | -0.039541727 |
| PredCluster4 | -0.057008659 |

### Standard Model and Hierarchical Model for CHF

The following table shows C-statistics for the Standard Model and the Hierarchical Model.

Standard Model and Hierarchical Model for CHF The following table shows C-statistics for the Standard Model and the Hierarchical Model.
|  | C-Statistic | CI95 (Min) | CI95 (Max) |
| --- | --- | --- | --- |
| Standard Model | 0.600 | 0.595 | 0.605 |
| Hierarchical Model | 0.600 | 0.595 | 0.606 |

The following table shows C-statistics for the Standard Model used to predict readmission for patients in each bicluster separately.

|  | C-Statistic | CI95 (Min) | CI95 (Max) |
| --- | --- | --- | --- |
| Bicluster 1 | 0.570 | 0.560 | 0.580 |
| Bicluster 2 | 0.587 | 0.576 | 0.598 |
| Bicluster 3 | 0.597 | 0.585 | 0.608 |
| Bicluster 4 | 0.614 | 0.602 | 0.625 |

### CMS Models (CMS Standard Model and CMS Hierarchical Model) for CHF

The following table shows C-statistics for the CMS Standard Model and the CMS Hierarchical Model.

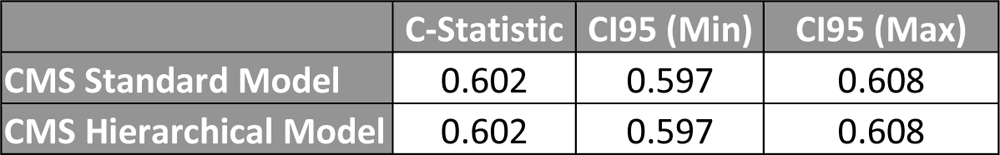

CMS Models (CMS Standard Model and CMS Hierarchical Model) for CHF The following table shows C-statistics for the CMS Standard Model and the CMS Hierarchical Model.
|  | C-Statistic | CI95 (Min) | CI95 (Max) |
| --- | --- | --- | --- |
| CMS Standard Model | 0.602 | 0.597 | 0.608 |
| CMS Hierarchical Model | 0.602 | 0.597 | 0.608 |

The following table shows C-statistics for the CMS Standard Model used to predict readmission for patients in each bicluster separately.

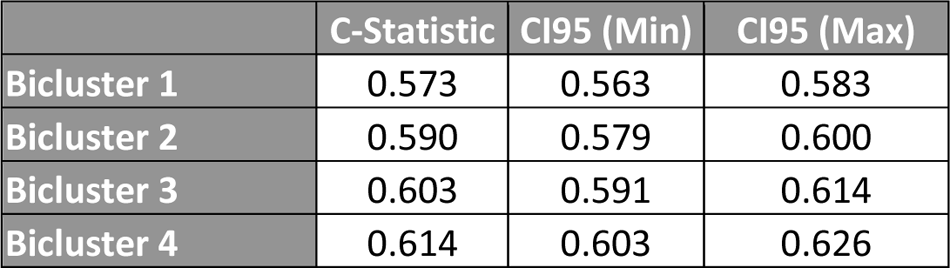

|  | C-Statistic | CI95 (Min) | CI95 (Max) |
| --- | --- | --- | --- |
| Bicluster 1 | 0.573 | 0.563 | 0.583 |
| Bicluster 2 | 0.590 | 0.579 | 0.600 |
| Bicluster 3 | 0.603 | 0.591 | 0.614 |
| Bicluster 4 | 0.614 | 0.603 | 0.626 |

### Model Coefficients for CMS Standard Model (CHF) [15]

|  |  |
| --- | --- |
| (Intercept) | -2.062052423 |
| V1 | -0.008807384 |
| V2 | 0.08069205 |
| V3 | 0.026511089 |
| V4 | 0.059292935 |
| V5 | 0.021486724 |
| V6 | 0.083551993 |
| V7 | 0.043335937 |
| V8 | 0.110433094 |
| V9 | 0.050644153 |
| V10 | 0.055051891 |
| V11 | 0.063148831 |
| V12 | 0.046434593 |
| V13 | 0.130099365 |
| V14 | -0.001401658 |
| V15 | 0.096204102 |
| V16 | 0.074628886 |
| V17 | 0.02554376 |
| V18 | 0.045885408 |
| V19 | 0.072014832 |
| V20 | 0.053421316 |
| V21 | 0.124207717 |
| V22 | 0.105487347 |
| V23 | 0.051450424 |
| V24 | 0.021372813 |
| V25 | 0.01075549 |
| V26 | 0.086498498 |
| V27 | -0.001335018 |
| V28 | 0.070577466 |
| V29 | 0.142129454 |
| V30 | 0.046599464 |
| V31 | 0.042236327 |
| V32 | 0.07795193 |
| V33 | 0.206227519 |
| V34 | 0.170726009 |
| V35 | 0.05424802 |
| V36 | 0.071097061 |
| V37 | 0.117969103 |

### Model Coefficients for CMS Hierarchical Model (CHF)

|  |  |
| --- | --- |
| (Intercept) | -2.050961429 |
| V1 | -0.008696382 |
| V2 | 0.080417206 |
| V3 | 0.024568048 |
| V4 | 0.060401182 |
| V5 | 0.026470131 |
| V6 | 0.07824069 |
| V7 | 0.055357171 |
| V8 | 0.100353589 |
| V9 | 0.061970384 |
| V10 | 0.066594667 |
| V11 | 0.068254076 |
| V12 | 0.058624423 |
| V13 | 0.120852698 |
| V14 | 0.005596376 |
| V15 | 0.102047312 |
| V16 | 0.079661612 |
| V17 | 0.031011406 |
| V18 | 0.050815364 |
| V19 | 0.084441452 |
| V20 | 0.065024208 |
| V21 | 0.119511648 |
| V22 | 0.11654221 |
| V23 | 0.047832743 |
| V24 | 0.0189412 |
| V25 | 0.007449149 |
| V26 | 0.097154811 |
| V27 | 0.009679744 |
| V28 | 0.079037213 |
| V29 | 0.147713008 |
| V30 | 0.04662239 |
| V31 | 0.041108317 |
| V32 | 0.088384596 |
| V33 | 0.190553921 |
| V34 | 0.160246633 |
| V35 | 0.035498901 |
| V36 | 0.061096207 |
| V37 | 0.106604226 |
| PredCluster2 | 0.028721208 |
| PredCluster3 | -0.029432347 |
| PredCluster4 | -0.048839832 |

#### TKA/THA

##### Discrimination

The following are box plots of risk prediction by readmission status for the Standard Model and the Hierarchical Model.

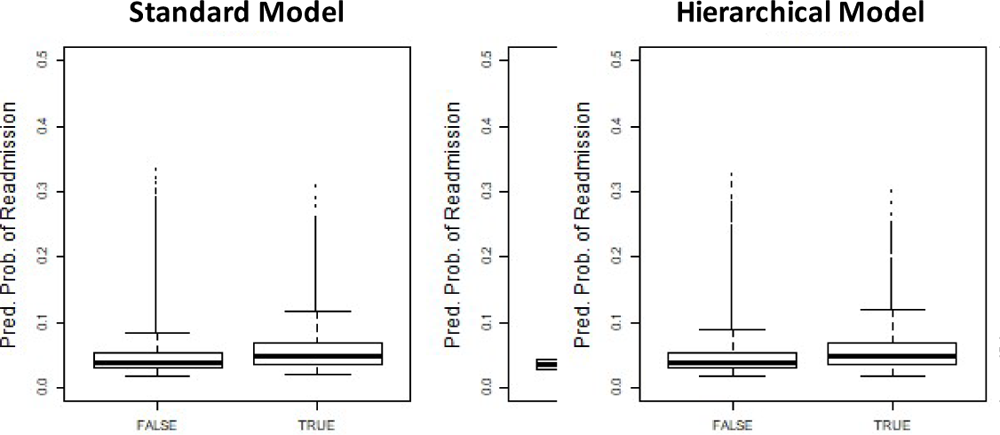

#### Calibration

The following are calibration plots showing logistic regressions relating readmission status to the logistic quantile of the predicted probability of readmission yield regression lines with specified intercept and slope (the ideal regression line would have intercept=0 and slope=1, which is shown shaded for reference). The histograms at the bottom of each graph reflect the frequency of modeled data, horizontal axes show predicted probability, vertical axes show actual probability, and the axes of all figures are constrained to range from 0 to .5.

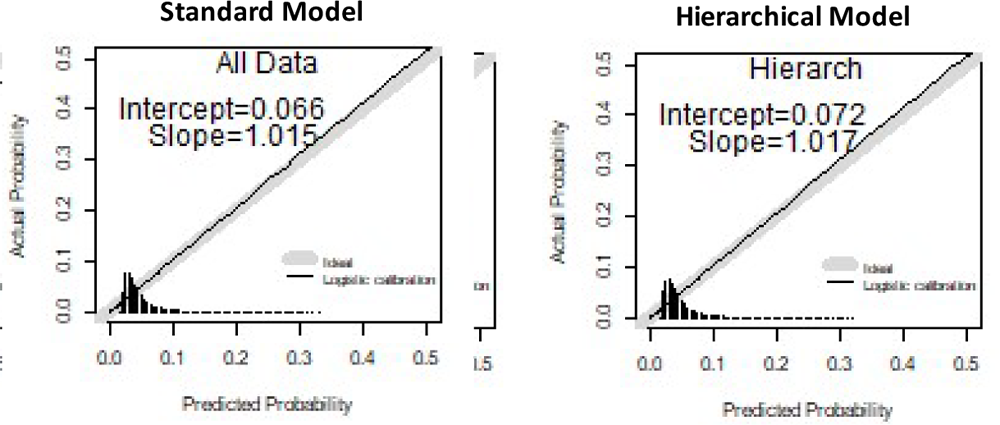

#### Coefficients

The following are logistic regression coefficients relating readmission status to the logistic quantile of the predicted probability of readmission for each model, with standard errors.

|  | Intercept (Standard Error) | Slope (Standard Error) |
| --- | --- | --- |
| Standard Model | 0.066 (0.092) | 1.015 (0.031) |
| Hierarchical Model | 0.072 (0.092) | 1.017 (0.031) |

### Model Coefficients for Standard Model (TKA/THA)

|  |  |
| --- | --- |
| (Intercept) | -6.263520554 |
| sex2 | -0.156977542 |
| AgeAdm | 0.037561114 |
| RACE51 | 0.107946335 |
| RACE52 | 0.385560747 |
| RACE53 | 0.083620721 |
| RACE55 | 0.103221123 |
| Renal_failure | 0.367756519 |
| Major_Symp_Abnormalities | 0.173001919 |
| Hypertension_Uncomp | 0.115321908 |
| Hypertension_Comp | 0.034487102 |
| Morbid_OB | 0.1284828 |
| Endocrine_disorders | 0.221828594 |
| Psychiatric_disorders | 0.321401406 |
| CHF | 0.21488135 |
| Coronary_angina | 0.263264479 |
| Arrhythmia | 0.24838679 |
| COPD | 0.389660791 |

### Model Coefficients for Hierarchical Model (TKA/THA)

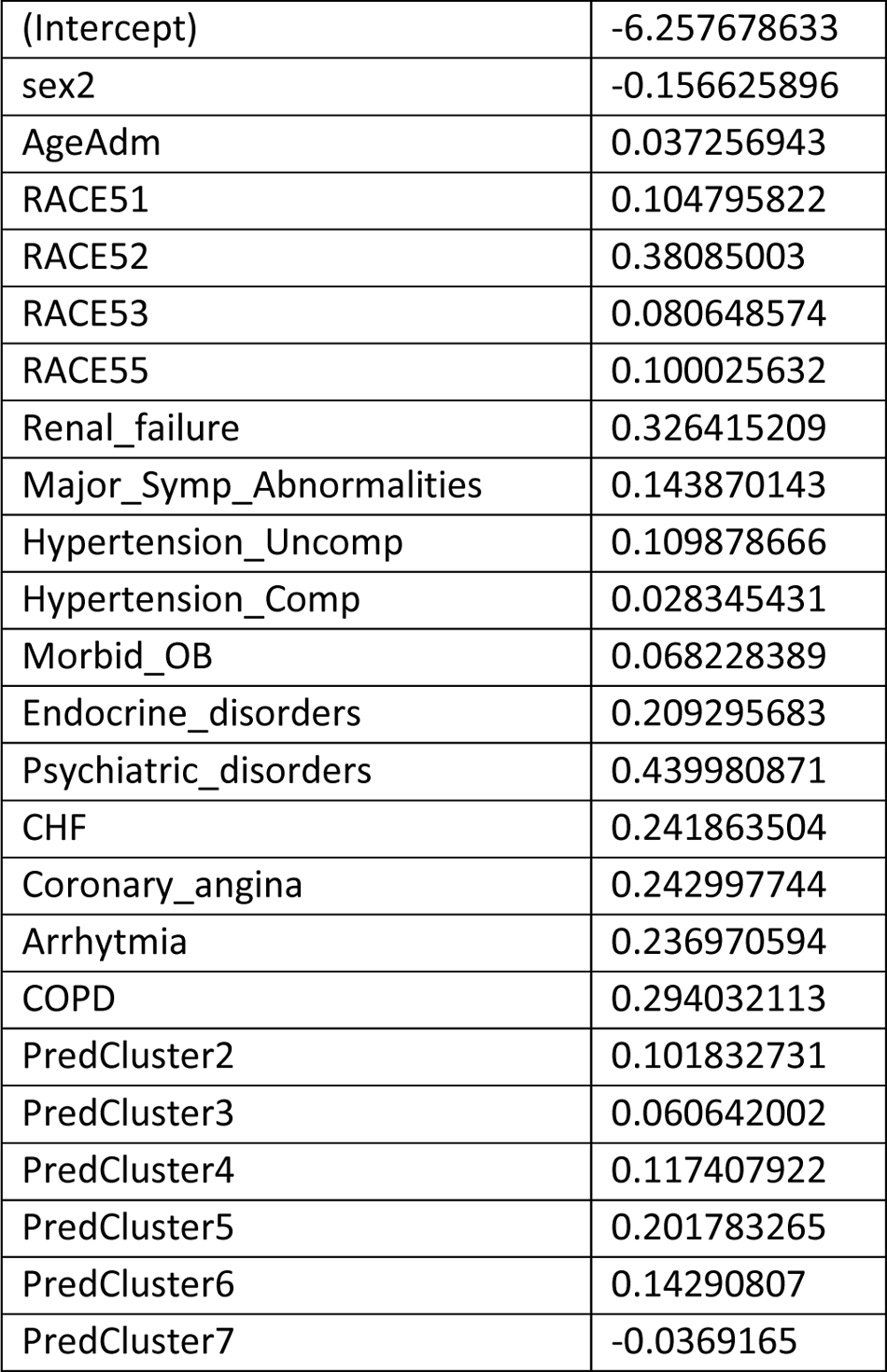

|  |  |
| --- | --- |
| (Intercept) | -6.257678633 |
| sex2 | -0.156625896 |
| AgeAdm | 0.037256943 |
| RACE51 | 0.104795822 |
| RACE52 | 0.38085003 |
| RACE53 | 0.080648574 |
| RACE55 | 0.100025632 |
| Renal_failure | 0.326415209 |
| Major_Symp_Abnormalities | 0.143870143 |
| Hypertension_Uncomp | 0.109878666 |
| Hypertension_Comp | 0.028345431 |
| Morbid_OB | 0.068228389 |
| Endocrine_disorders | 0.209295683 |
| Psychiatric_disorders | 0.439980871 |
| CHF | 0.241863504 |
| Coronary_angina | 0.242997744 |
| Arrhythmia | 0.236970594 |
| COPD | 0.294032113 |
| PredCluster2 | 0.101832731 |
| PredCluster3 | 0.060642002 |
| PredCluster4 | 0.117407922 |
| PredCluster5 | 0.201783265 |
| PredCluster6 | 0.14290807 |
| PredCluster7 | -0.0369165 |

### Standard Model and Hierarchical Model for THA/TKA

The following table shows C-statistics for the Standard Model and the Hierarchical Model.

Standard Model and Hierarchical Model for THA/TKA The following table shows C-statistics for the Standard Model and the Hierarchical Model.
|  | C-Statistic | CI95 (Min) | CI95 (Max) |
| --- | --- | --- | --- |
| Standard Model | 0.638 | 0.629 | 0.646 |
| Hierarchical Model | 0.638 | 0.629 | 0.647 |

The following table shows C-statistics for the Standard Model used to predict readmission for patients in each bicluster separately.

|  | C-Statistic | CI95 (Min) | CI95 (Max) |
| --- | --- | --- | --- |
| Bicluster 1 | 0.578 | 0.553 | 0.603 |
| Bicluster 2 | 0.608 | 0.584 | 0.631 |
| Bicluster 3 | 0.595 | 0.570 | 0.620 |
| Bicluster 4 | 0.610 | 0.585 | 0.635 |
| Bicluster 5 | 0.584 | 0.559 | 0.609 |
| Bicluster 6 | 0.579 | 0.553 | 0.606 |
| Bicluster 7 | 0.636 | 0.617 | 0.655 |

### CMS Models (CMS Standard Model and CMS Hierarchical Model) for TKA/THA

#### C-Statistics

The following table shows C-statistics for the CMS Standard Model and the CMS Hierarchical Model.

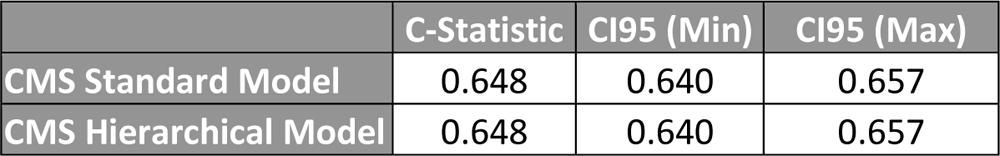

The following table shows C-statistics for the CMS Standard Model used to predict readmission for patients in each bicluster separately.

|  | C-Statistic | CI95 (Min) | CI95 (Max) |
| --- | --- | --- | --- |
| Bicluster 1 | 0.585 | 0.561 | 0.609 |
| Bicluster 2 | 0.632 | 0.609 | 0.655 |
| Bicluster 3 | 0.617 | 0.594 | 0.641 |
| Bicluster 4 | 0.630 | 0.606 | 0.653 |
| Bicluster 5 | 0.608 | 0.583 | 0.632 |
| Bicluster 6 | 0.589 | 0.562 | 0.615 |
| Bicluster 7 | 0.640 | 0.621 | 0.659 |

### Model Coefficients for CMS Standard Model (TKA/THA) [16]

|  |  |
| --- | --- |
| (Intercept) | -4.007625537 |
| Var1 | 0.036039292 |
| Var2 | -0.149437339 |
| Var3 | 0.088157516 |
| Var4 | 0.252326647 |
| Var5 | -0.255229777 |
| Var6 | 0.234030925 |
| Var7 | 0.131248955 |
| Var8 | 0.118415513 |
| Var9 | -0.014234681 |
| Var10 | 0.158442117 |
| Var11 | 0.186149829 |
| Var12 | 0.10822119 |
| Var13 | 0.120553114 |
| Var14 | 0.213179137 |
| Var15 | 0.166404922 |
| Var16 | 0.244882944 |
| Var17 | 0.260963406 |
| Var18 | 0.228775313 |
| Var19 | 0.099773803 |
| Var20 | 0.167375189 |
| Var21 | 0.236389437 |
| Var22 | 0.08572084 |
| Var23 | 0.22805813 |
| Var24 | 0.092669018 |
| Var25 | 0.099767352 |
| Var26 | 0.363700358 |
| Var27 | 0.038962561 |
| Var28 | 0.582409201 |
| Var29 | 0.317306501 |
| Var30 | 0.103490531 |
| Var31 | -0.001329201 |
| Var32 | 0.026795269 |
| Var33 | 0.150796704 |

### Model Coefficients for CMS Hierarchical Model (TKA/THA)

|  |  |
| --- | --- |
| (Intercept) | -4.0316433 |
| Var1 | 0.035679265 |
| Var2 | -0.148849333 |
| Var3 | 0.08807725 |
| Var4 | 0.25290998 |
| Var5 | -0.257380043 |
| Var6 | 0.241117802 |
| Var7 | 0.132427427 |
| Var8 | 0.115796146 |
| Var9 | -0.014769398 |
| Var10 | 0.15814836 |
| Var11 | 0.187159646 |
| Var12 | 0.035368612 |
| Var13 | 0.120886777 |
| Var14 | 0.211303719 |
| Var15 | 0.170049309 |
| Var16 | 0.245655958 |
| Var17 | 0.387470462 |
| Var18 | 0.234387939 |
| Var19 | 0.102192659 |
| Var20 | 0.187162267 |
| Var21 | 0.20830413 |
| Var22 | 0.084196245 |
| Var23 | 0.206829049 |
| Var24 | 0.096990789 |
| Var25 | 0.103278456 |
| Var26 | 0.256166878 |
| Var27 | 0.069134573 |
| Var28 | 0.601609967 |
| Var29 | 0.294460582 |
| Var30 | 0.109616297 |
| Var31 | 0.003492241 |
| Var32 | 0.030762491 |
| Var33 | 0.117221489 |
| PredCluster2 | 0.132189893 |
| PredCluster3 | 0.074260901 |
| PredCluster4 | 0.096335076 |
| PredCluster5 | 0.225685704 |
| PredCluster6 | 0.166772439 |
| PredCluster7 | -0.034196246 |

